# Machine Learning Identifies Microbiome and Clinical Predictors of Sustained Weight Loss Following Prolonged Fasting

**DOI:** 10.1101/2025.06.26.25330331

**Authors:** Gelsomina N. Kaufhold, Theda U. P. Bartolomaeus, Kristin Kräker, Till Schütte, Sakshi Kamboj, Ulrike Löber, Gabriele Rahn, Victoria McParland, Lena Braun, Lajos Markó, Matanat Mammadli, Alexander Krannich, Lina S. Bahr, Friederike Gutmann, Friedemann Paul, Quinten R. Ducarmon, Georg Zeller, Robin Mesnage, Nicola Wilck, Alma Zernecke, Peter J. Oefner, Wolfram Gronwald, Dominik N. Müller, Sofia K. Forslund-Startceva, Sylvia Bähring, Hendrik Bartolomaeus, Nadja Siebert

**Affiliations:** Experimental and Clinical Research Center (ECRC), a cooperation of Charité - Universitätsmedizin Berlin and Max Delbrück Center for Molecular Medicine, Berlin, Germany; Charité-Universitätsmedizin Berlin, corporate member of Freie Universität Berlin and Humboldt-Universität zu Berlin, Berlin, Germany; Max Delbrück Center for Molecular Medicine in the Helmholtz Association (MDC), Berlin, Germany; DZHK (German Centre for Cardiovascular Research), partner site Berlin, Germany; Centre for Ecological and Evolutionary Synthesis (CEES), Department of Biosciences, University of Oslo, Oslo, Norway; Institute of Functional Genomics, University of Regensburg, Regensburg, Germany; Department of Nephrology and Internal Intensive Care Medicine, Charité –Universitätsmedizin Berlin, Germany; BioStats GmbH, Nauen, Germany; Molecular Systems Biology Unit, European Molecular Biology Laboratory (EMBL), Heidelberg, Germany; Leiden University Center for Infectious Diseases (LUCID), Leiden University Medical Center (LUMC), Leiden, Netherlands; Science Department, Buchinger Wilhelmi Clinic, Überlingen, Germany; Department of Nutritional Sciences, School of Life Course Sciences, Faculty of Life Sciences and Medicine, King’s College London, London, United Kingdom; Institute of Experimental Biomedicine, University Hospital Würzburg, Germany

**Keywords:** fasting, gut microbiome, weight loss, machine learning, personalized nutrition, precision medicine, multi-omics, prevention, metabolic health

## Abstract

**Background:** Prolonged fasting may improve metabolic health, but controlled data in healthy adults with longer follow-up and multi-omics profiling are limited. We investigated short- and long-term effects of a 5-day fasting intervention on body composition, gut microbiome, and circulating and fecal metabolites, and assessed whether baseline characteristics predict individual weight-loss response.

**Methods:** In a randomized, waitlist-controlled trial, 38 healthy adults completed a 5-day fasting intervention with 12-week follow-up (LEANER study). Outcomes included body mass index and body composition, gut microbiome composition, and plasma and fecal metabolites. Changes over time and between groups were evaluated using regression-based models and paired non-parametric tests, as appropriate. Additionally, permutation-based multivariate testing was performed on microbiome and metabolome data. Twelve-week body weight response was predicted using data-driven machine learning with cross-validation, followed by external validation in three independent cohorts undergoing prolonged fasting protocols.

**Results:** Fasting reduced body mass index acutely, predominantly driven by loss of fat mass, and these improvements partially persisted at 12 weeks. Fasting induced marked shifts in gut microbiome composition and in plasma and fecal metabolites. Post-fasting and longer-term changes in microbial diversity were associated with baseline microbiome diversity. A model combining baseline microbiome and clinical variables predicted body mass index response at 12 weeks; prominent predictors included an unclassified *Faecalibacterium* species, *Oscillibacter* sp. 50_27, low-density lipoprotein cholesterol, and systolic blood pressure. The model generalized to three independent cohorts, including individuals with metabolic syndrome, patients with multiple sclerosis exposed to repeated fasting, and healthy volunteers fasting for 6–12 days.

**Conclusions:** In healthy adults, a 5-day prolonged fasting intervention produces robust short-term metabolic changes with partial persistence and consistent remodeling of the gut microbiome and metabolite profiles. Baseline microbiome and clinical characteristics can help stratify expected longer-term responses, supporting the development of individualized fasting-based interventions.

**Trial registration:** ClinicalTrials.gov, NCT04452916. Registered on June 29, 2020

## Introduction

Fasting interventions have gained renewed attention for their potential to modulate metabolism, reshape the gut microbiome, and promote healthy aging^1^. While fasting protocols vary widely from time-restricted eating to fasting-mimicking diets^2^, and prolonged food abstinence lasting up to several weeks (up to 26 days)^3^, the underlying principle is consistent: transient caloric deprivation triggers coordinated physiological responses that may confer long-term health benefits^4^.

Evidence from patient populations, including those with metabolic syndrome^5^, autoimmune diseases^6,7^, and cancer^8^, suggests that fasting can influence key regulatory pathways such as autophagy, insulin sensitivity, and oxidative stress^1^. In individuals with metabolic syndrome, we previously demonstrated that a 5-day fast atop a DASH (Dietary Approaches to Stop Hypertension) diet induced profound and coordinated changes in gut microbiome composition and immune cell states most of which were reversed at 12 weeks follow-up^5^. Using immune cell population or microbiome profiles, we developed models that predicted long-term antihypertensive effects, supporting the concept of data-driven stratification^5^. Metabolic effects of weight loss interventions such as fasting may be assessed by determination of resting energy expenditure (REE). REE is defined as the energy required to sustain vital physiological functions while at rest and without any external activity or food intake^9^. It accounts for approximately 60–75% of total daily energy expenditure and is the most significant contributor to overall metabolism^9^. The value is influenced by factors including age, sex, genetics, and body composition, with lean muscle mass being a dominant determinant^10^. Changes in REE are central to understanding metabolic adaptations and the effects of interventions, whether nutritional, pharmacological, or involving lifestyle changes. Reliable measurement is typically achieved through indirect calorimetry, considered the gold standard^11,12^. Changes in REE may reflect body responses to interventions and contribute to understanding individual variability in weight management and energy requirements.

In parallel, gut microbiome profiling has emerged as a powerful tool for personalized medicine^13^. Microbial signatures have been linked to energy metabolism^14^, weight regulation^15,16^, and immune modulation^17,18^. Personalized dietary approaches that account for microbiome composition have demonstrated superior outcomes in metabolic control^8^. Emerging applications extend to autoimmune, neuropsychiatric, and oncologic indications, underscoring the potential of microbiome-informed strategies in precision medicine^13,19^. Despite these advances, the long-term effects of prolonged fasting in healthy individuals remain poorly characterized. Most existing studies rely on non-randomized designs, lack adequate control groups, or fail to include follow-up assessments. Furthermore, many interventions are performed in multimodal clinical settings involving exercise, psychological support, and dietary coaching^5^, making it difficult to isolate the effect of fasting alone.

Here, we present a randomized, waitlist-controlled study investigating the effects of a 5-day prolonged fasting intervention in healthy adults, with follow-up assessments at 12 weeks. We integrated clinical, microbiome, and metabolome profiling to characterize immediate and long-term physiological responses. Using machine learning (ML), we show that sustained weight loss can be predicted from baseline microbiome and clinical features. Results were validated in one independent healthy cohort and cohorts with metabolic syndrome and multiple sclerosis. Our findings offer mechanistic insight into fasting responses and support the potential of microbiome-informed strategies for personalized lifestyle interventions.

## Results

### Prolonged fasting affects energy expenditure and long-term weight loss

Thirty-eight healthy participants were recruited for a 5-day prolonged fasting intervention (Figure 1a). As fasting interventions cannot be blinded, we decided to use a waiting list control to account for physiological fluctuations within a 12-week timeframe while maximizing the number of participants undergoing prolonged fasting (Figure 1A). Baseline characteristics are shown in Table 1.

**Table 1.** Baseline characteristics of healthy participants depending on their randomization to fasting and waiting control arm. Data are given as means (SD). p-values fasting vs. waiting by t-test. BMI, body mass index; eGFR, estimated glomerular filtration rate; ALT, alanine aminotransferase; TSH, thyroid-stimulating hormone; CRP, C-reactive protein; HbA1c, glycated hemoglobin.

| Variable | All | Fasting | Waiting | P value |
| --- | --- | --- | --- | --- |
| Number (n) | 38 | 19 | 19 |  |
| Sex, female | 19 / 38 (50%) | 9 / 19 (47%) | 10 / 19 (53%) | >0.9 |
| Age (years) | 38 (8) | 39 (9) | 36 (8) | 0.4 |
| BMI (kg/m<sup>2</sup>) | 25.1 (2.5) | 25.0 (2.7) | 25.2 (2.3) | 0.7 |
| eGFR<br>(ml/min/1.73m<sup>2</sup>) | 102 (14) | 98 (12) | 106 (15) | 0.1 |
| ALT (U/l) | 23.1 (8.1) | 23.0 (8.2) | 23.2 (8.2) | 0.2 |
| TSH (mU/l) | 1.70 (0.84) | 1.77 (0.91) | 1.64 (0.77) | 0.6 |
| CRP (mg/dl) | 0.98 (0.87) | 0.78 (0.28) | 1.17 (1.17) | 0.2 |
| Cholesterol (mg/dl) | 182 (36) | 189 (35) | 175 (36) | 0.2 |
| HbA1c (mmol/mol) | 33.2 (2.8) | 34.2 (2.6) | 32.3 (2.7) | 0.03 |
| Hemoglobin (g/dl) | 14.0 (1.3) | 13.8 (1.3) | 14.2 (1.2) | 0.41 |
| White blood count<br>(uL<sup>-1</sup>) | 5.46 (1.02) | 5.56 (0.96) | 5.37 (1.09) | 0.6 |
| Thrombocytes (uL<sup>-1</sup>) | 240 (42) | 238 (63) | 239 (54) | 0.9 |

**Figure 1.**
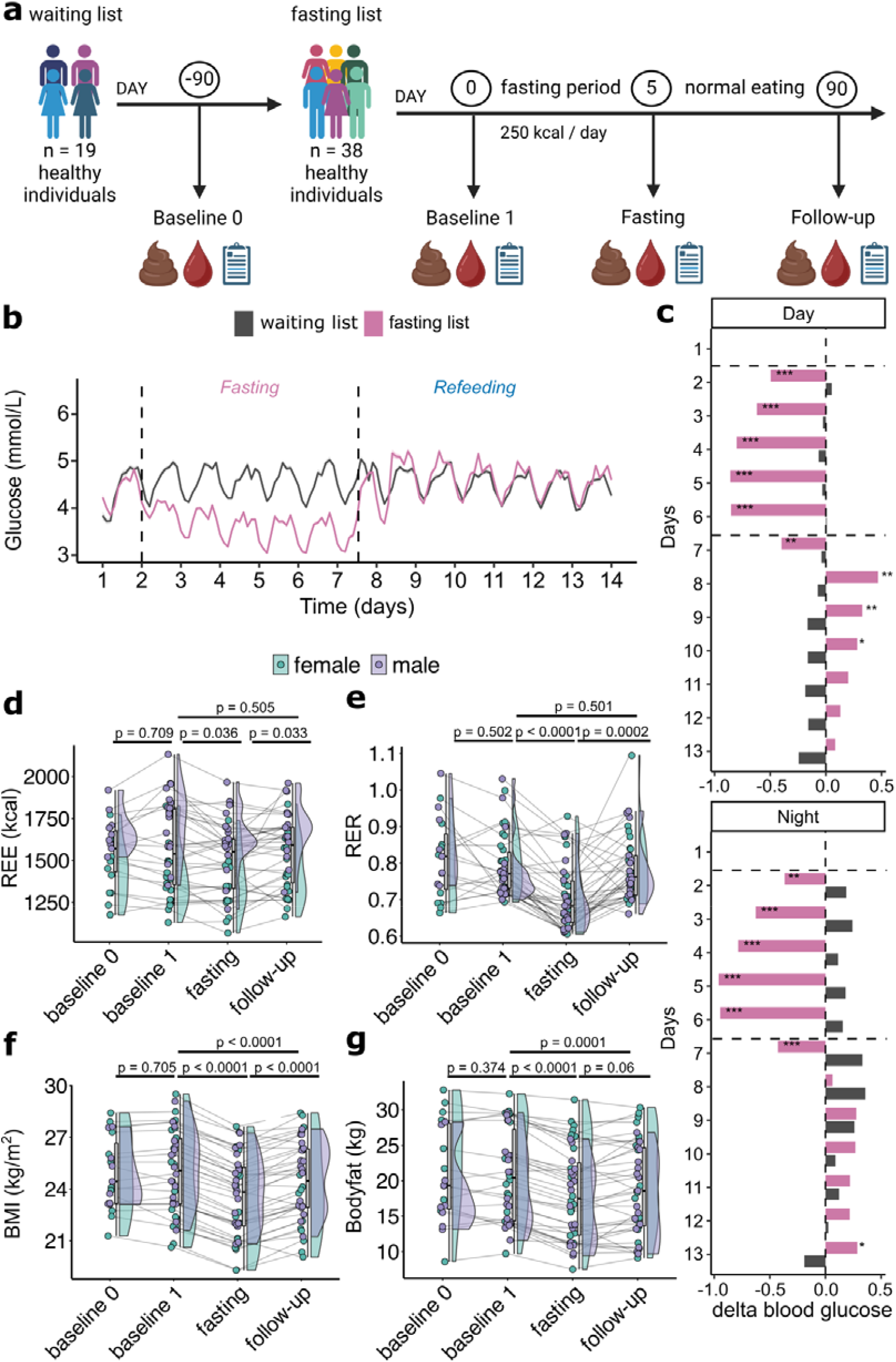
Physiological effects of prolonged fasting in healthy individuals. **a)** Schematic overview of the randomized waitlist-controlled trial. A total of 38 healthy participants were recruited. The first 19 participants immediately underwent prolonged fasting, while the remaining 19 served as a waitlist control group. After the initial intervention period, the waitlist group also underwent the same prolonged fasting protocol, resulting in a complete dataset of 38 participants undergoing prolonged fasting. The fasting intervention consisted of a 5-day period with 250 kcal/day, preceded and followed by baseline and follow-up visits at days 0, 5, and 90. Stool, blood, and clinical data were collected at each visit. The waitlist group was assessed at an additional baseline (day -90) to control for temporal fluctuations. **b)** Glucose time series measured by continuous intradermal monitoring starting one day prior to fasting (pink) or at baseline for the waiting list control (grey). **c)** Quantification of absolute changes in mmol/L of mean daytime (6am to 10pm) (top) and night (10pm to 6am) glucose levels (bottom), with changes compared to the individual baseline (day 1). Paired t-test with Benjamini-Hochberg false discovery rate correction (BH-FDR), *q<0.1, **q < 0.01, ***q < 0.001. **d)** Resting energy expenditure (REE), **e)** respiratory exchange ratio (RER), **f)** body mass index (BMI), and **g)** body fat percentage at the different study visits. Each point represents one participant, lines connect individuals, boxplot (middle) for all participants, violins (right side) and dots coloured by sex. P-values are from Wilcoxon signed-rank tests and were corrected using the Holm method.

Of the 38 participants, 83% reported fasting-related adverse events (AE) (Table S1). All AE were mild and transient and did not necessitate discontinuation of fasting. Blood glucose and β-hydroxybutyrate (BHB) levels were measured each morning and evening during the five-day fasting period (days 1–5), confirming that all participants adhered closely to the study protocol (Figure S1a, b). We observed a diurnal rhythm with higher BHB but lower glucose concentrations in the morning compared to the evening (Figure S1a, b). Continuous interstitial glucose monitoring during the five-day fasting period, compared with baseline measurements and waiting-list controls, confirmed a dynamic reduction in glucose levels over the course of the fasting period (Figure 1b). Following refeeding, the glucose levels normalized; however, between days 2 and 4 post-fast, blood glucose concentrations were elevated throughout the day (06:00–22:00) compared with baseline measurements (Figure 1c).

Our predefined endpoint was a change in REE. Prolonged fasting acutely reduced REE by 62 kcal/d, suggesting a fasting-induced, energy-preserving mechanism (Figure 1d, Supplementary Data 1). We did not observe alterations in REE between the two baseline timepoints of waiting list controls (Figure 1d). Following the findings of earlier work^10^, we performed a linear mixed model showing that lean mass (p < 0.001) and not self-reported sex significantly influenced REE at all visits, including post-fasting (Figure S2). Following the five-day fast, respiratory exchange ratios (RER) declined significantly (Figure 1e), paralleling a peak in BHB concentrations on day 5 (Figure S1b). All those metabolic markers returned to baseline by 12 weeks post-fast.

Furthermore, we observed that prolonged fasting reduced BMI (Figure 1f). This decrease was partly reversed at the 12-week follow-up (Figure 1f). Body composition analysis attributed this incomplete return towards baseline BMI predominantly to a reduction in fat mass (Figure 1g, S1g). Of note, the absolute and relative reduction of body fat was accompanied by a reduction in lean mass during fasting that completely reverted at follow-up (Figure S1h). Prolonged fasting reduced waist-to-hip ratio and diastolic blood pressure while increasing heart rate, findings which are consistent with a previously published study^5^ in metabolic syndrome (Figure S1d-g). In contrast to metabolic syndrome patients^5^, the healthy participants in this study did not exhibit sustained blood-pressure reduction (Figure S1e, f). Taken together, these results demonstrate that prolonged fasting is well tolerated in healthy individuals, yielding sustained reductions in BMI and fat mass, while most metabolic and hemodynamic parameters return to baseline during the 12-week follow-up.

### Prolonged fasting induces large short-term and small long-term changes in gut microbiome composition

To assess the natural fluctuation of the gut microbiome composition, we analyzed changes between the two baseline samples (baseline 0 and baseline 1) from the waiting list control (Figure 1a). Microbial alpha diversity, measured as Shannon index, and microbiome composition quantified by Bray-Curtis dissimilarity did not differ significantly between the two baseline samples (Figure S3a, b). Shannon index remained stable across all study visits (Figure 2a). In contrast, individual trajectories in response to fasting varied markedly (Figure 2a). Interestingly, short-term (post-fasting) and long-term (follow-up) changes in alpha-diversity correlated negatively with alpha-diversity at baseline, implicating baseline differences in microbiome composition as relevant predictors of fasting-induced microbiome changes (Figure 2b, c). Bray-Curtis dissimilarity-based multivariate analysis of the taxonomic composition demonstrated a significant shift in microbial community composition in response to fasting that partially reversed during the follow-up period (Figure 2d). Consistent with the observed shift in the multivariate analysis, fasting induced significant changes in several bacterial species that reversed during follow-up. For example, *Roseburia sp*. inserta sedis and *Bifidobacterium adolescentis* decreased after the fasting period but increased at follow-up, whereas *Faecalibacterium sp*. CAG-74 showed the opposite pattern (Figure 2e, S3c, Supplementary Data 2). In contrast, only seven taxa exhibited sustained abundance changes (Figure S3c).

**Figure 2.**
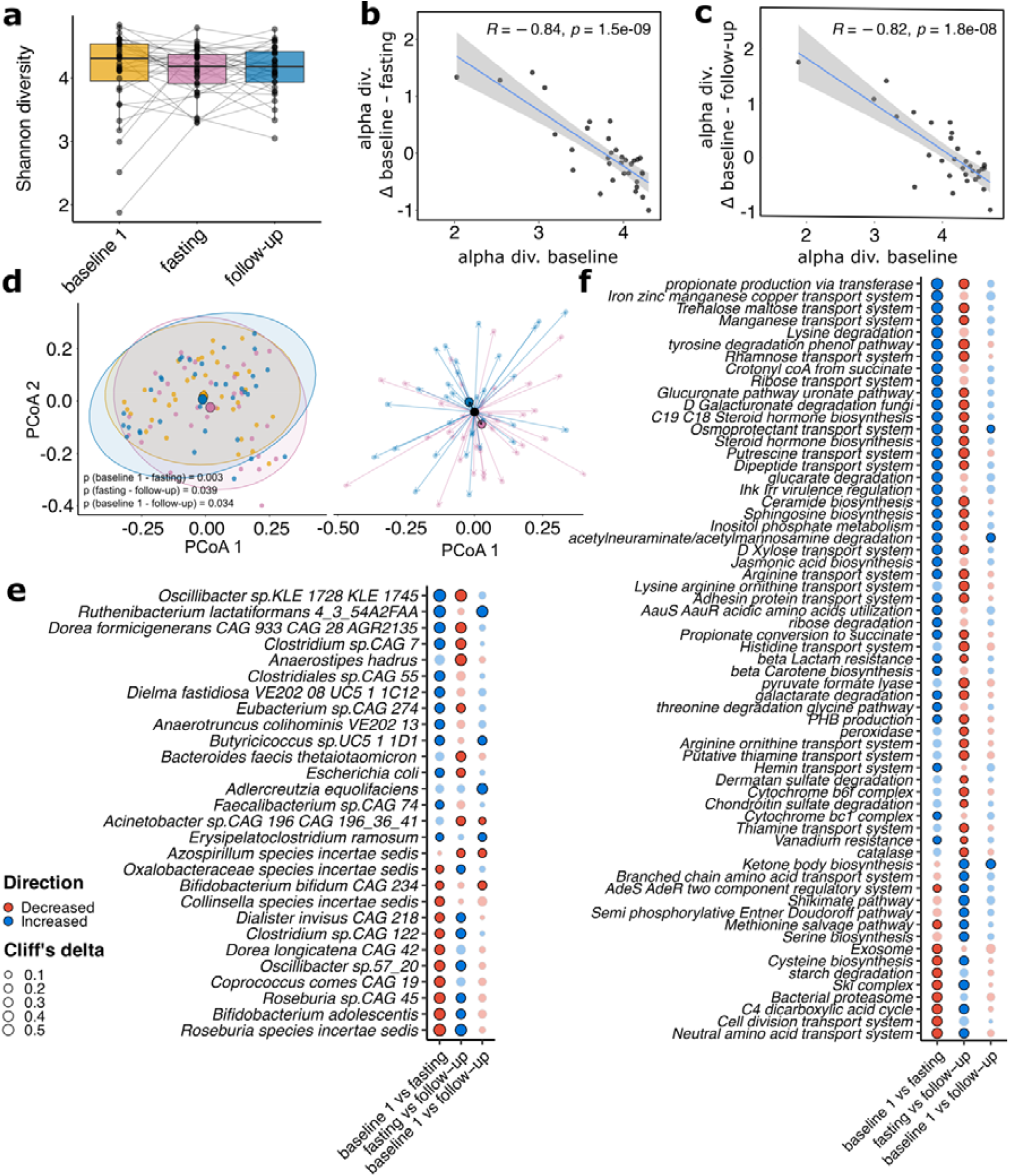
Prolonged fasting induces transient alterations in gut microbiome composition and function. **a)** Alpha diversity as measured by Shannon index for the study visits. Correlation of Shannon index at baseline with changes in Shannon index immediately **b)** post-fasting and **c)** at 12-week follow-up, indicating that baseline diversity predicts microbiome responsiveness. **d)** Principal coordinate analysis (PCoA) based on Bray–Curtis dissimilarity (naïve, left) and centered on baseline visit for fasting effect and on fasting visit for post-fasting effect (right) show a transient shift in overall microbial composition after fasting (pink), which partially reverts at follow-up (blue). Ellipses represent 95% confidence intervals, p-values from PERMANOVA. Dot plots showing differential abundant e) species (taxonomic level) and f) gut metabolic modules (functional level). Dots show fasting effects, follow-up effects and study effects, transparency indicates non-significant findings (q>0.1), dot size shows absolute Cliff’s delta, color shows directionality. Significance by Wilcoxon signed-rank test with Benjamini–Hochberg false discovery rate correction.

Next, we analyzed gut-specific metabolic modules using GOmixer^20^. Again, fasting induced pronounced alterations that largely reverted to baseline during follow-up (Figure 2f). Only for three modules, ketone body synthesis, osmoprotectant transport system, and N-acetylneuraminate and N-acetylmannosamine degradation, fasting induced a long-lasting increase when comparing baseline to follow-up (Figure 2f, Figure S4a, Supplementary Data 3).

Overall, prolonged fasting induces pronounced compositional shifts in the gut microbiome, which were largely reversed upon refeeding, resulting in minimal long-term deviations from baseline at the 12-week follow-up. Notably, baseline alpha diversity metrics correlate with the magnitude of alpha diversity changes both immediately after fasting and at follow-up.

### Prolonged fasting alters plasma and fecal metabolome

We performed nuclear magnetic resonance (NMR) spectroscopy on plasma and fecal samples to assess the impact of fasting, and its associated microbiome shifts on the fecal and host metabolome. Principal coordinate analysis (PCA), centered on baseline values to characterize post-fast changes and on post-fast values to characterize follow-up effects, revealed plasma metabolome trajectories analogous to those observed in the gut microbiome (Figure S4b). In line, we observed significant alterations between baseline, fasting, and follow-up, with most changes reversed at follow-up (Figure S4c, Supplementary Data 4). In plasma, only valine, glycine, tyrosine, and pyruvic acid exhibited sustained increases from baseline to follow-up, indicating lasting effects of fasting on these metabolites (Figure S4b, c).

Fecal metabolite profiling again revealed a pattern of alteration and reversion to baseline (Figure S4d). Contradicting other previous reports on short-chain fatty acids (SCFA) in fasting^5^, we observed a decrease in butyrate during fasting, which was non-significant at follow-up (Figure S4e, Supplementary Data 5). The only long-term effect observed in our data was a decrease in choline from baseline to fasting (Figure S4e).

In conclusion, NMR-based metabolomics of plasma shows more consistent alterations as compared to the fecal metabolome, which might be due to the different matrix or intraindividual variations in gut microbiome composition.

### Prediction of sustained BMI response by multi-omics machine learning

Since we observed strong intraindividual differences in response to prolonged fasting across most data layers, we aimed to investigate whether the clinical response to fasting can be predicted from baseline data. BMI showed the overall strongest long-term effect of the analyzed clinical data (Figure 1). Therefore, we focused on the long-term weight loss response as the primary phenotype that we aimed to predict. Using the waiting list control, we estimated the normal fluctuation of BMI within 12 weeks of study participation. Normal fluctuation in BMI was defined as mean ± two standard deviations (SD)^21^, in our case -0.008 ± 0.32 kg/m^2^. Based on this threshold, we identified 38% long-term responders and 62% non-responders to prolonged fasting.

To predict BMI loss response, a random forest classifier was trained on 87 features including the 50 most abundant microbial species at baseline, additional candidate taxa and pathways from previous prolonged fasting studies, and relevant clinical metadata (Supplementary Data 6). To obtain a model that would be as transferable as possible to publicly available datasets, we focused on microbiome and clinical features rather than NMR metabolomics^5,22^. Due to the small sample size, we applied a repeated cross-validation approach to ensure robust model evaluation. Specifically, we used 100 repeated stratified 80/20 resamples, each incorporating an inner 5-fold cross-validation loop for hyperparameter tuning. Model performance was further optimized by performing feature selection once after model tuning, based on random-forest feature importance, the Boruta algorithm, and backward feature elimination (Figure 3b, S5c). This resulted in four features: LDL, systolic blood pressure (BP), *Faecalibacterium sp*. incerta sedis and *Oscillibacter sp*. 57_20. Those four features were used to train the final model. Across the repeated 80/20 resampling, the refined short model performed well with an AUC of 0.84 ± 0.17 (Table 2).

**Table 2.** Summary of predictive performance metrics for the predefined sparse 4-feature model, evaluated using 100 iterations of stratified 80/20 Monte Carlo resampling. Values are reported as mean ± SD across resamples. AUC, area under the receiver operating characteristic curve; F1, F1 score (harmonic mean of precision and recall); MCC, Matthews correlation coefficient; SD, standard deviation.

| Metric | Value (mean ± SD) |
| --- | --- |
| AUC | 0.84 ± 0.17 |
| Accuracy | 0.76 ± 0.15 |
| Sensitivity | 0.56 ± 0.34 |
| Specificity | 0.85 ± 0.17 |
| Balanced Accuracy | 0.71 ± 0.18 |
| F1 | 0.68 ± 0.18 |
| MCC | 0.50 ± 0.35 |

**Figure 3.**
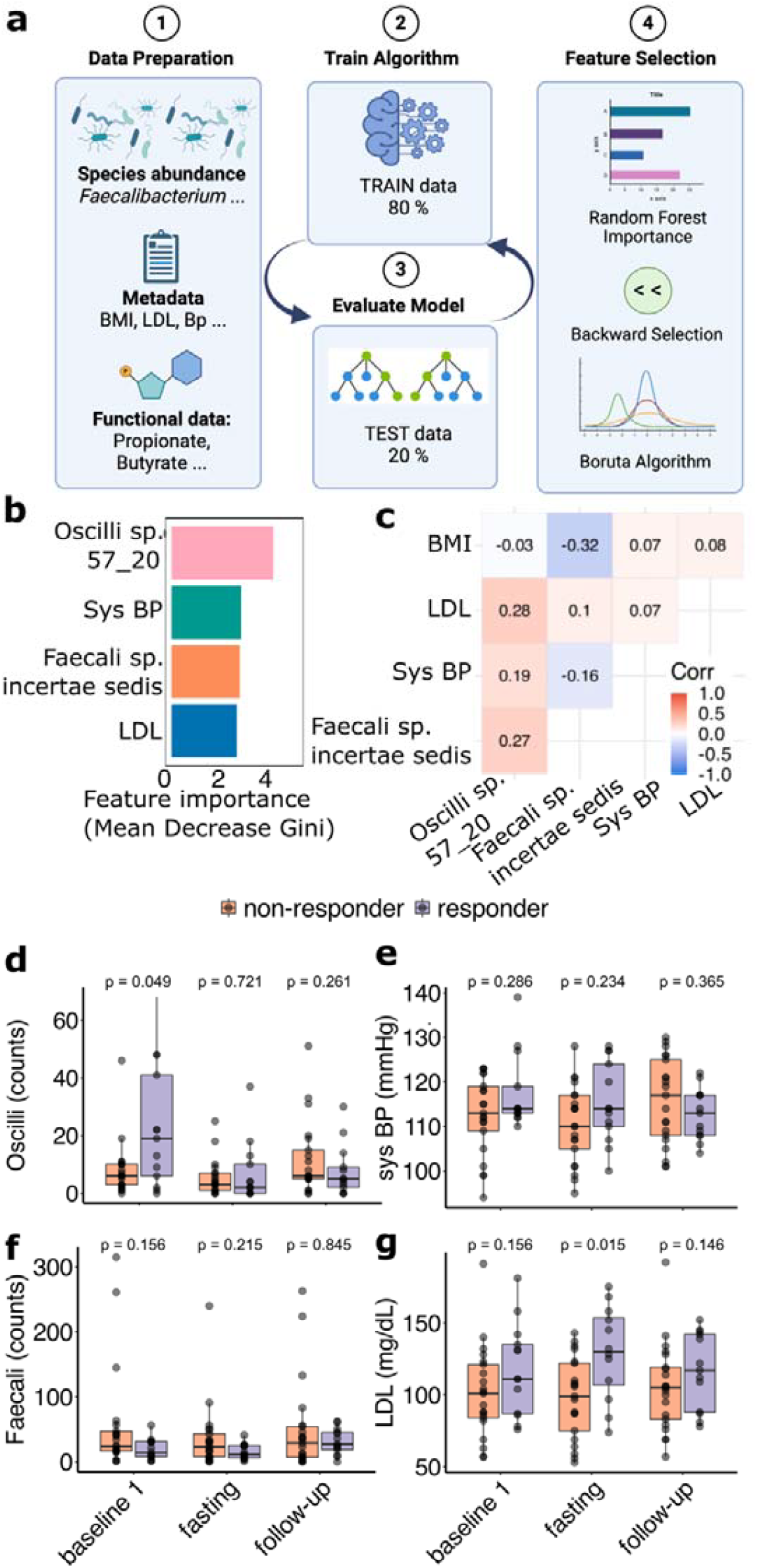
Machine learning model predicts sustained BMI response at follow-up to prolonged fasting using clinical and microbiome features. **a)** Schematic overview of the multi*-omics* feature selection and classification pipeline. Baseline species abundances, clinical metadata, and functional modules were used to train a random forest classifier. Important features were selected through recursive feature elimination, Boruta algorithm, and backward selection. **b)** Final model includes four top-ranked features: *Faecalibacterium* sp. incertae sedis, *Oscillibacter* sp. 57_20, LDL cholesterol, and systolic blood pressure. **c)** Correlation heatmap of the selected features showing independence from baseline BMI. R-values from Spearman correlation. **d)** Boxplots of the four selected features at baseline, comparing BMI responders and non-responders, dots show individual patients. P-values by Mann-Whitney-U-test. Oscilli, *Oscillibacter* sp. 57_20; Faecali, *Faecalibacterium* sp. incertae sedis; LDL, Low-density lipoprotein cholesterol; sys BP, systolic blood pressure.

Baseline BMI can confound weight loss prediction. Therefore, we tested collinearity between baseline BMI and our four selected features, none of which reached significance (Figure 3c). To test if the four selected features are mere surrogate markers of baseline BMI, we incorporated baseline BMI into the model. This did not improve predictive accuracy (Figure S5b). Furthermore, *Oscillibacter sp*. 57_20 remained the most important feature. Feature-stability analysis supported this ranking: *Oscillibacter sp*. 57_20 and *Faecalibacterium sp*. incertae sedis consistently ranked highest, followed by systolic BP, while LDL showed low and inconsistent importance (−0.9 ± 3.2; Supplementary Table S2a). To avoid post-hoc feature removal, LDL was retained in the sparse model, although a 3-feature variant performed similarly (Supplementary Table S3). Next, we compared a clinical-only model (including baseline BMI, blood pressure, and lipid parameters) with a combined model that additionally incorporated microbiome data to address potential confounding by clinical features (Table 3). The clinical-only model achieved an AUC = 0.69 ± 0.21, accuracy = 0.66 ± 0.18, and balanced accuracy = 0.62 ± 0.19 (Table 3). The combined model improved performance across all metrics (AUC = 0.86 ± 0.17, accuracy = 0.74 ± 0.16, balanced accuracy = 0.72 ± 0.18, Table 3). These results indicate that microbiome features contribute predictive information beyond standard clinical covariates, underscoring their added value for identifying responders to prolonged fasting.

**Table 3.** Comparison of predictive performance of clinical-only versus clinical+microbiome models. Predictive performance of models trained on clinical features alone compared with models including both clinical and microbiome features. Values represent mean ± SD across 100 repeated stratified 80/20 resamples. Inclusion of microbiome data improved classification metrics across all performance measures, indicating additional predictive value beyond clinical covariates. AUC, area under the receiver operating characteristic curve; SD, standard deviation; F1, F1-score; MCC, Matthews correlation coefficient.

| **Metric** | **Clinical-only model** | **Clinical + Microbiome model** |
| --- | --- | --- |
| AUC | 0.69 ± 0.21 | 0.86 ± 0.17 |
| Accuracy | 0.66 ± 0.18 | 0.74 ± 0.16 |
| Sensitivity | 0.52 ± 0.34 | 0.66 ± 0.35 |
| Specificity | 0.73 ± 0.22 | 0.78 ± 0.19 |
| Balanced Accuracy | 0.62 ± 0.19 | 0.72 ± 0.18 |
| F1 | 0.60 ± 0.18 | 0.69 ± 0.16 |
| MCC | 0.28 ± 0.41 | 0.48 ± 0.37 |

Finally, we examined the abundance patterns of the four features from the reduced model within our cohort. Responders exhibited significantly higher levels of *Oscillibacter sp*. 57_20 (Figure 3d), while the identified *Faecalibacterium sp*. showed no significant difference (Figure 3f). Although systolic BP and LDL contributed to model classification, their individual baseline levels did not differ significantly between responders and non-responders (Figure 3e, g). Taken together, we identified four (two clinical and two microbiome) features that jointly hold the potential to predict long-term weight loss in response to prolonged fasting intervention. Although performance remained consistently above random across repeated cross-validation, we emphasize the expected variability due to small cohort size and interpret the findings as exploratory but reproducible evidence of predictive potential.

### Application of the classifier in independent cohorts

To test transferability in independent datasets, we applied the final model to data from three publicly available cohorts that differed in health status and fasting design. Given these differences, these analyses are treated as exploratory applications rather than formal external validations.

As a first step, we applied our model to our published metabolic syndrome cohort, in which patients underwent either a five-day fasting regimen followed by a DASH diet or a DASH diet alone (Figure 4a, n=34)^5^. The study outcomes were assessed 12 weeks post-intervention, mirroring our study’s timeline. Given the greater BMI variability expected in metabolic syndrome patients, we quantified BMI fluctuation from the DASH only group (control group). Long-term responders were again defined as exceeding the mean plus two standard deviation weight loss of the control group (DASH only), corresponding to BMI reduction exceeding 0.8 kg/m^2^. Remarkably, our trained model predicted sustained weight loss in this study with an AUC of 0.85 (Figure 4b-c, Supplementary Table S3). Next, we calibrated the model using Platt scaling and isotonic regression. Despite stable strong discrimination, the Brier score remained unchanged after calibration (Supplementary Table S3), indicating that residual error likely reflects differences in outcome prevalence or underlying data distributions rather than miscalibration alone. Consistent with our healthy-participant cohort, the model’s predictive features exhibited only weak correlations with baseline BMI, underscoring their independence from initial body mass (Fig. 4d). In univariate analyses, *Oscillibacter sp*. 50_27 and *Faecalibacterium sp*. showed significant differences at baseline between responders and non-responders, while clinical features showed no difference (Figure 4e-h).

**Figure 4.**
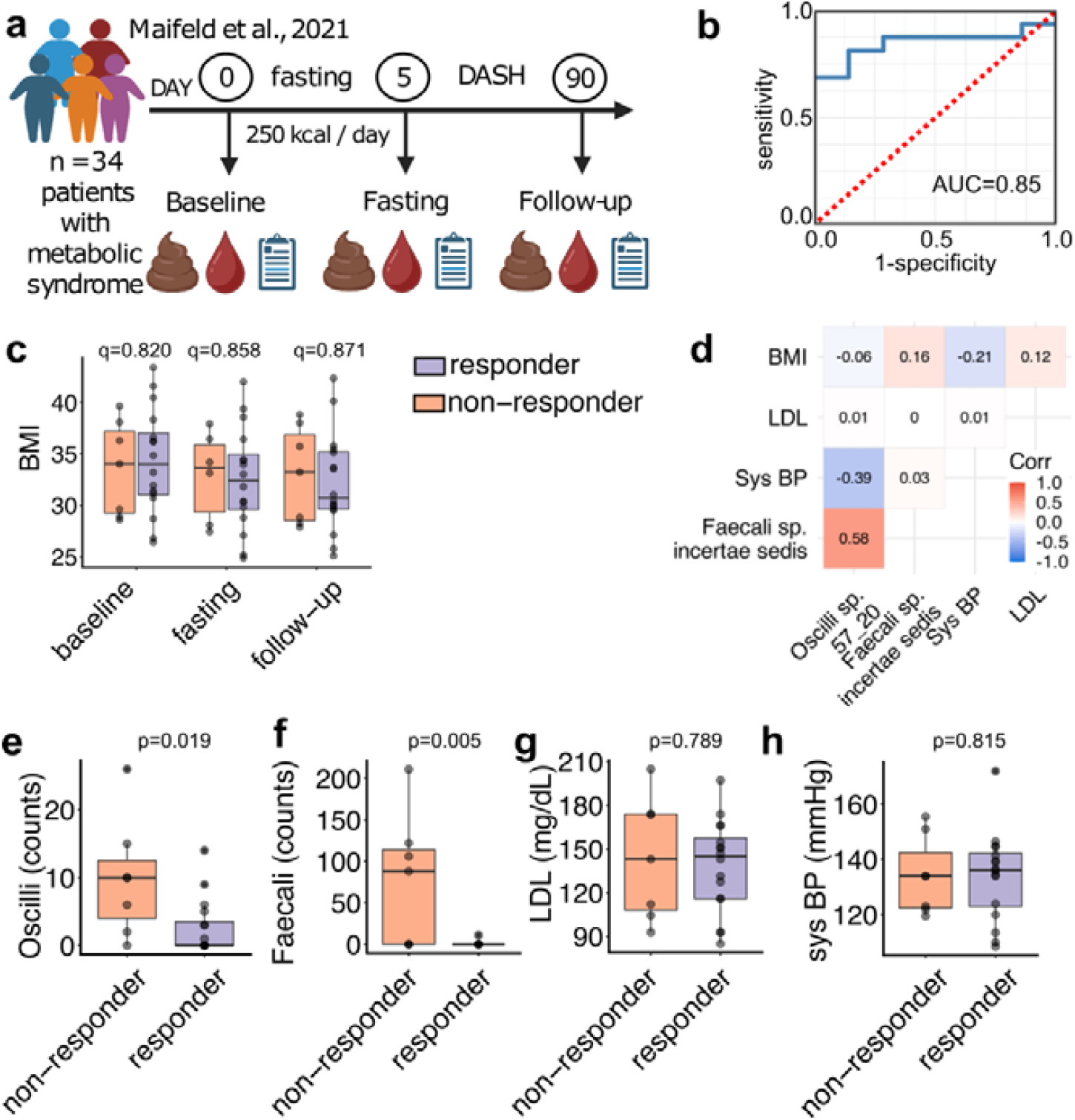
Application of BMI response prediction model in an external fasting cohort of patients with metabolic syndrome. **a)** Schematic of the Maifeld et al. (2021) study: 5-day fasting followed by a DASH diet in individuals with metabolic syndrome (n = 34), with BMI follow-up after 12 weeks. **b)** Receiver operating characteristic (ROC) curve of model performance in the external Maifeld cohort, showing good predictive accuracy (AUC = 0.85). **c)** BMI over the course of the study, split by participants classified as responders or non-responders. Each point represents a participant. P-values are from paired Wilcoxon signed-rank tests and were corrected for multiple comparisons using the Benjamini-Hochberg false discovery rate (FDR-BH) method. **d)** Correlation heatmap of model features with BMI. Features used for classification at baseline: **e)** *Oscillibacter* sp. 57_20, f) *Faecalibacterium* sp. incertae sedis, g) LDL, and h) systolic blood pressure. Oscilli, *Oscillibacter* sp. 57_20; Faecali, *Faecalibacterium sp*. incertae sedis; LDL, Low-density lipoprotein cholesterol; sys BP, systolic blood pressure.

As a further dataset, we applied the classifier to a recent nutritional intervention trial in patients with multiple sclerosis, the majority of whom exhibited normal metabolic profiles (obesity in one patient, hypertension in two, and elevated LDL in four)^22^. Notably, the study design differed from the other cohorts. Patients completed two prolonged 7-day periods of fasting (Figure 5a, n=22). However, these prolonged fasting periods were interspersed with intermittent fasting (14 hrs/ day), and follow-up assessments were conducted nine months apart (Figure 6a). Because no control arm was available to isolate the effects of the prolonged fasting periods from those of intermittent fasting, we could not define average BMI variation and thus compute specificity and sensitivity or perform calibration. However, patients classified as responders by our model experienced significantly greater weight loss than those classified as non-responders (Figure 5b). To provide an alternative quantitative assessment, we evaluated the correlation between the probability of responder classification and the observed magnitude of BMI change (Figure 5c). While the Spearman correlation did not achieve statistical significance (p=0.08), the direction and magnitude of the association (ρ=0.309) support the model’s ability to reflect observed BMI changes. No significant correlations between model parameters and baseline BMI were detected (Figure 5d). Similar to the current cohort, levels of *Oscillibacter sp*. 50_27 were significantly higher in responders (Figure 5e), while the other features did not differ significantly (Figure 5f-h).

**Figure 5.**
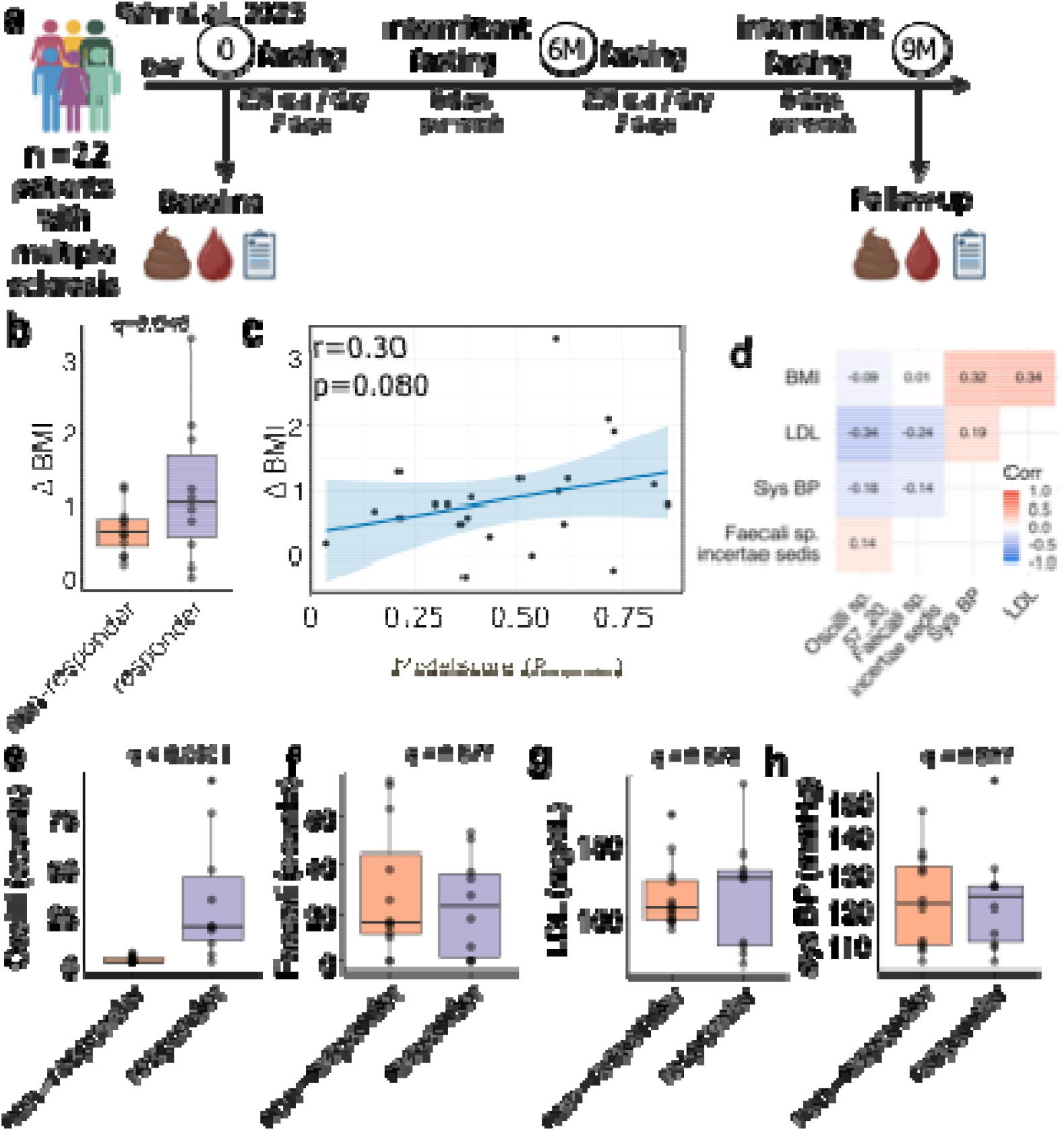
Application of BMI response prediction model in an external fasting cohort of patients with multiple sclerosis. **a)** Schematic of the Bahr et al. (2025) study: two 7-day periods of prolonged fasting and intermittent fasting in individuals with multiple sclerosis (n = 22), over a 9-month period. **b)** BMI reduction at follow-up of patients classified as responder or non-responder by the prediction model. p-value by one-tailed unpaired t-test. **c)** Correlation analysis of changes in BMI with probability for responder status derived from the model prediction. P-value by one-tailed Spearman correlation. **d)** Correlation heatmap of model features with BMI. Features used for classification at baseline: ***e)*** *Oscillibacter* sp. 57_20, ***f)*** *Faecalibacterium* sp. incertae sedis, **g)** LDL, and g) systolic blood pressure. Oscilli, *Oscillibacter* sp. 57_20; Faecali, *Faecalibacterium* sp. incertae sedis; LDL, Low-density lipoprotein cholesterol; sys BP, systolic blood pressure.

**Figure 6.**
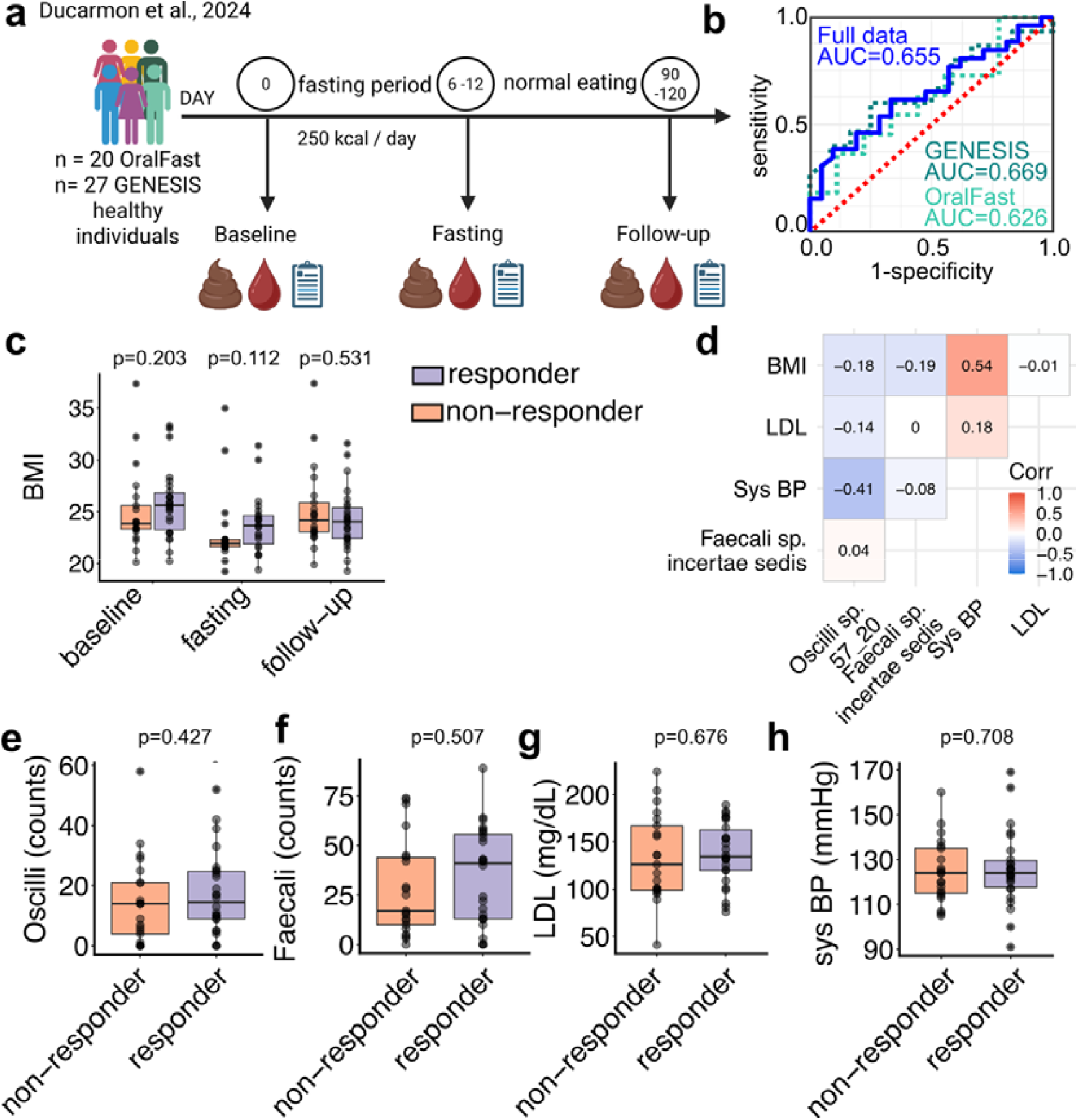
Application of BMI response prediction model in an external fasting cohort of healthy volunteers. **a)** Schematic of the Ducarmon et al. (2024) study: 6-12-days prolonged fasting with BMI follow-up after 3 months in the OralFast subcohort (n = 20) or 4 months in the GENESIS subcohort (n=27). **b)** Receiver operating characteristic (ROC) curve of model performance showing intermediate accuracy (AUC = 0.655). **c)** BMI over the course of the study, split by participants classified as responders or non-responders. Each point represents a participant. P-values are from paired Wilcoxon signed-rank tests and were corrected for multiple comparisons using the Benjamini-Hochberg false discovery rate (FDR-BH) method. **d)** Correlation heatmap of model features with BMI. Features used for classification at baseline: ***e)*** *Oscillibacter* sp. 57_20, ***f)*** *Faecalibacterium* sp. incertae sedis, **g)** LDL, and h) systolic blood pressure. Oscilli, *Oscillibacter* sp. 57_20; Faecali, *Faecalibacterium* sp. incertae sedis; LDL, Low-density lipoprotein cholesterol; sys BP, systolic blood pressure.

Lastly, we applied the model to available data from healthy volunteers undergoing between 6 and 12 days of prolonged fasting^23^. This dataset comprises two subcohorts: the GENESIS (ClinicalTrials.gov: NCT05449249) and OralFast (ClinicalTrials.gov: NCT05031598) trials. Both follow similar study designs (Figure 6a) but differ in their follow-up durations — 3 months for OralFast and 4 months for GENESIS. Outcome was assessed after 16 weeks during a study visit and as self-reported outcome after 12 weeks, respectively (Figure 6a, n=47). Since this study was performed in healthy participants, we used the same BMI cut-offs for responder definition as used for the model training in our cohort. Our trained model predicted sustained weight loss in this study with an AUC of 0.655. (Figure 6b-c, Supplementary Table S4). Of note, AUC was slightly better for the GENESIS than the OralFast data (Figure 6b). Platt scaling and isotonic regression did not improve model calibration but retained consistent discrimination (Supplementary Table S4). In contrast to the other cohorts, baseline BMI and systolic blood pressure were correlated (Fig. 6d). In univariate analyses, none of the features differed significantly at baseline between responders and non-responders (Figure 6e h).

Our findings indicate that baseline patient characteristics determine the magnitude of long-term weight response to prolonged fasting interventions. Furthermore, the predictive model, initially developed in healthy subjects, was successfully applied to independent cohorts investigating prolonged fasting. However, given the differences in health status, study design, and the absence of an intermittent fasting-only group in the multiple sclerosis cohort, this represents an exploratory rather than a formal validation. The observed association between predicted responder status and actual BMI reduction provides supportive but not confirmatory evidence of external applicability, highlighting the need for larger, harmonized fasting studies to confirm generalizability.

*Metabolite associations with Oscillibacter sp*. 57_20 *and Faecalibacterium sp*. incertae sedis Since both the *Oscillibacter sp*. 57_20 and the *Faecalibacterium sp*. identified in the LEANER were predictive of weight loss, we wanted to further understand the potential metabolite profile associated with these bacteria. Therefore, we performed an exploratory correlation analysis with the fecal metabolome across all visit data while blocking the visit information^24^. We found significant associations of *Faecalibacterium sp*. with butyrate, tryptophan, and uridine (Figure 7a-c, Supplementary Data 7) as well as of *Oscillibacter sp*. 57_20 with glycerol and methionine, and for uridine without reaching statistical significance (Figure 7d-f, Supplementary Data 8). In summary, our data suggests that the bacterial species important for the classification of long-term body weight responders modulate the metabolomic output of the fecal microbiome.

**Figure 7.**
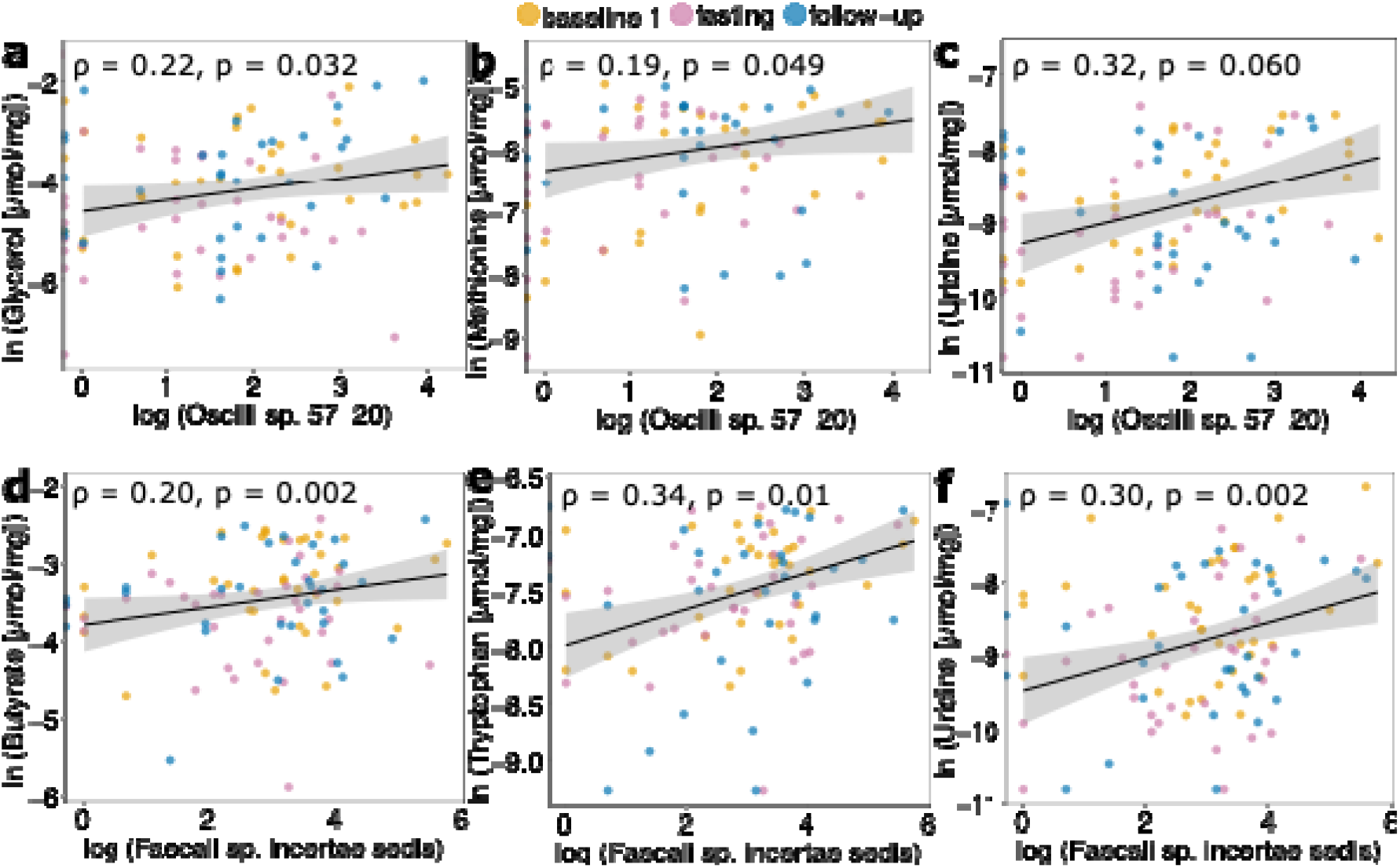
*Oscillibacter* sp. 57_20 and *Faecalibacterium* sp. incertae sedis abundance correlates with fecal metabolome. Correlation of *Oscillibacter* (Oscilli) sp. 57_20 abundance with fecal **a)** glycerol, **b)** methionine, and **c)** uridine levels. Correlation of *Faecalibacterium* (Faecali) sp. incertae sedis abundance with fecal **d)** butyrate, **e)** tryptophan, and **f)** uridine levels. R-values from Spearman’s correlation. P-values from permutation-based independence test screening for associations of species with metabolites while blocking effects of study visits. Abbreviations: Oscilli: *Oscillibacter* sp. 57_20; Faecali: *Faecalibacterium* sp. incertae sedis.

## Discussion

Our data show that prolonged fasting leads to long-term body weight and body fat reduction after 12 weeks. Interindividual variability in response to dietary interventions poses a major challenge in lifestyle interventions. Large-scale studies have shown that both host and microbial factors shape individual metabolic responses, emphasizing the need for precision strategies^25,26^. Using baseline microbiome and clinical data, we can predict individuals who will respond to fasting with sustained weight loss, defined as a BMI reduction lower than the mean minus two standard deviations of the waiting list control. Using available data from similar interventions in healthy volunteers, patients with metabolic syndrome, and patients with multiple sclerosis, we could apply our data-driven classification approach. While *Faecalibacterium sp*. incertae sedis and *Oscillibacter sp*. 57_20, were important for BMI response prediction, most of the microbiome and metabolome features were characterized by transient shifts induced by fasting, with subsequent return to pre-intervention states. Taken together, our data implicate gut microbiota in long-term BMI changes post fasting and suggest conserved effects of fasting regardless of health status. We used a data-driven definition of responders, defining weight loss beyond the mean plus two standard deviations of the control group in each cohort (the waiting-list control in the current study and the DASH-only arm in the Maifeld et al. cohort). This represents a statistical rather than a clinical threshold, chosen to capture changes exceeding normal physiological variation^21^. While this approach ensures objectivity across cohorts, it may not fully reflect clinically meaningful improvements, which future studies with larger and harmonized samples should define more precisely.

On the host level, prolonged fasting led to strong changes in multiple cardiometabolic risk factors like LDL cholesterol, waist-to-hip ratio, and systolic and diastolic blood pressure. However, at the 12-week follow-up these changes had mostly reverted, indicating that prolonged fasting did not exert a sustained effect in healthy individuals. These data differ from patients with metabolic syndrome with values above the healthy range^5^. Interestingly, while blood glucose dropped during fasting, it increased upon refeeding during the day (6am to 10pm). Since values at night (10pm to 6am) did not increase, our data suggest a mildly disturbed glucose handling for the early days post-fasting. A similar phenotype was reported in the literature in response to standardized meal intakes after prolonged fasting^27,28^. This was attributed to the metabolism successfully switching to fatty acid and ketone oxidation and postprandial carbohydrate oxidation was reduced, leading to higher plasma glucose levels^27^. Since a strong metabolic phenotype was expected, REE reduction was our predefined endpoint. REE in fasting was studied previously mainly in healthy men^29-31^. Our data shows an overall similar but smaller effect, which might be due to the inclusion of healthy women and a broader range of lean masses. REE reduction after 24h caloric deprivation was previously associated with weight gain after 6 months in healthy individuals^32^. We could not confirm these findings in our cohort for long-term weight changes after 3 months using the changes in REE after 5 days of fasting (Figure S6). However, using a combination of clinical and microbiome data, we were successful in predicting long-term weight loss.

We have shown in the past the microbiome and immune data can predict blood pressure reduction in patients with metabolic syndrome in response to fasting^5^. In contrast, our study in healthy participants revealed only modest effects on blood pressure. Given the well-established impact of fasting on weight management^33^, we applied a data-driven approach that involved clustering participants based on BMI changes and employing a random forest classifier. Our initial model, which incorporated 83 features, demonstrated high sensitivity but limited specificity. Iterative feature selection enabled us to identify *Faecalibacterium sp*. incertae sedis, *Oscillibacter sp*. 57_20, plasma LDL levels, and systolic blood pressure for a feature-reduced classifier. Notably, the successful application of the model in independent cohorts highlighted the robustness and generalizability of our approach. In the Maifeld et al. cohort^5^, which featured a similar study design in patients with metabolic syndrome, we observed an 82.6% of responders were correctly identified. In the Bahr et al. cohort^22^, with a different study design incorporating intermittent fasting in patients with multiple sclerosis, predicted responders exhibited higher weight loss. Furthermore, we tested the model in a cohort of healthy volunteers published by Ducarmon et al.^23^. In this cohort, discrimination was moderate (AUC = 0.655), which we interpret as evidence of partial but attenuated signal transfer. Several methodological and biological factors likely contributed to this reduction in performance. First, the follow-up timing at 3 months for one subcohort and 4 months for the other differed from the standardized 12-week follow-up applied in both our primary study and the Maifeld et al. cohort. The longer interval increases the influence of post-intervention behavior, which weakens the link between baseline microbiota and long-term weight dynamics. Second, outcomes in the OralFast subcohort were self-reported rather than measured, introducing non-differential measurement error and potential bias. Self-reported weight is known to systematically underestimate true body weight change, particularly in non-obese individuals^34^, and such noise disproportionately affects probabilistic classifiers by compressing the outcome distribution. Third, the fasting protocol in Ducarmon et al. involved substantially deeper caloric restriction than the 5-day intervention used in our study. Longer fasting typically produces a larger initial weight loss driven by acute depletion of glycogen stores and associated water and lean mass changes^35^, thereby diluting the proportion of weight loss attributable to microbiota-dependent mechanisms. Furthermore, the fasting duration in itself was heterogenous among participants (6-12 days) and not standardized to a certain number of days^5,22^. Finally, the physiological measurements across the studies were not fully harmonized. Blood pressure, a feature that contributes to the clinical context of the model, was assessed in our study and in the Maifeld and Bahr cohorts using a standardized protocol with an oscillometric device (Mobil-O-Graph)^5,22^. In contrast, Ducarmon et al.^23^, used a different measurement approach, which may introduce variability in clinical covariates relative to the trained model. Notably, in this dataset, baseline BMI and systolic blood pressure were correlated, unlike in the other cohorts, suggesting population-specific interactions between anthropometric and cardiovascular traits that were not present during model training.

Taken together, these differences in study design, outcome measurement, physiological assessment, and fasting intensity likely account for the attenuated but still meaningful predictive performance in the Ducarmon dataset. The moderate AUC indicates that the classifier captures a biological signal that remains detectable across heterogeneous fasting contexts, but that true external validation will require cohorts with harmonized protocols, standardized follow-up durations, and objectively measured outcomes.

To assess whether calibration could improve probability estimates, we applied post-hoc calibration using Platt scaling and isotonic regression. However, these adjustments enhanced predictive performance in neither of the two external datasets. Post-hoc calibration often fails when the underlying problem is a mismatch between the training and validation cohorts rather than statistical miscalibration^36^. In our case, differences in fasting protocol, follow-up period, and even methodological aspects such as systolic blood pressure measurement, as mentioned above, likely contributed to the reduced model transferability. Calibration methods can only improve prediction when the target data share similar distributions and measurement contexts with the training data; they cannot correct for biological, behavioral, or procedural heterogeneity^37^. In contrast, the Maifeld et al. cohort demonstrated robust discrimination, but most likely the limited sample size constrained formal calibration. Together, these findings highlight that while our model retains general discriminative capacity across independent datasets, its probabilistic calibration and transportability depend on harmonized study protocols and consistent data collection practices.

These external explorations not only reinforce the reliability of our predictive model but also suggest that the gut microbiome changes associated with fasting responses are conserved across diverse populations, from healthy individuals to those with metabolic disorders and autoimmune disease, thereby broadening the potential applicability of fasting-based interventions for weight management and metabolic health.

An important aspect of our findings is the distinction between the multivariate classifier and univariate analyses of individual features. While none of the selected predictors were consistently significant when tested individually, their combined predictive value was clearly demonstrated by the random forest classifier^38^. This reflects the interdependent nature of biological systems, where linear and nonlinear relationships among variables often provide stronger discriminative power than single-feature associations^39^. However, univariate analyses remain valuable for hypothesis generation and mechanistic interpretation, as performed extensively across cohorts to contextualize individual predictors^39^. This pattern has also been observed in previous microbiome-based prediction studies, where integrative models outperform single-feature analyses by capturing interdependent microbial and host effects^40^.

At the same time, we recognize that differences in underlying disease, medication use, and dietary context across cohorts may confound microbiome–BMI associations and limit direct transferability of model coefficients^41^. The relatively small sample size of the training cohort adds further uncertainty, although consistent results in three independent datasets support the robustness of our approach. Therefore, the present model should be viewed as exploratory evidence that microbiome and clinical features together contain predictive information for fasting responses, rather than as definitive proof of generalizable biomarkers. Future validation in larger, prospectively harmonized fasting cohorts will be essential to confirm the robustness and clinical utility of these predictors.

Given that baseline BMI is typically a predictor of weight loss, we assessed its confounding effect on fasting-induced BMI changes. *Oscillibacter sp*. 57_20 remained the stronger predictor even when baseline BMI was included in the model, indicating that this microbial species may exert an independent influence on weight regulation during fasting. While we cannot fully exclude potential confounding or correlations between baseline BMI and microbiome composition, to our knowledge, no prior study has directly linked *Oscillibacter sp*. 57_20 with weight regulation. However, *Oscillibacter sp*. 57_20 was recently identified as a keystone taxon in a similar prolonged fasting study in healthy individuals, where it was implicated in the production of indole-3-propionate (I3P)^23^, a protective metabolite in cardiometabolic disease^42^. Future research is needed to explore the mechanistic role of *Oscillibacter spp*. in host metabolism and its potential relevance in personalized fasting interventions.

*Faecalibacterium spp*. are known SCFA producers^43^. In line, our data shows that the abundance of our specific *Faecalibacterium sp*. associates with fecal butyrate levels. As described previously for other *Faecalibacterium spp*.^*44*^, the identified *Faecalibacterium sp*. correlates with fecal tryptophan. Tryptophan can be catabolized by gut microbiota to indole metabolites^45^. Unfortunately, due to the resolution of NMR metabolomics, we could not detect specific indole metabolites like I3P. However, our data imply that both bacteria might interact in the production of tryptophan catabolites.

Both the *Faecalibacterium sp*. identified in this study and the *Oscillibacter sp*. 57_20 correlate with the nucleoside uridine. Uridine can adaptively regulate food intake in healthy participants^46^. Furthermore, experimental evidence show that blood uridine levels reflect the nutritional status of mice^47,48^ with high uridine levels as indicators of catabolic states^47,49^. In line, dietary supplementation of uridine enhanced hunger and increased caloric intake^46^. Microbial metabolism of uridine suggests a potential role of a gut-brain axis or influence on satiety^50^ in long-term BMI responders. Unfortunately, the literature concerning the other metabolites of interest correlating with *Oscillibacter sp*. 57_20, glycerol and methionine is sparse. While we controlled our analysis for abundance shifts at different visits, we could not control the false discovery rate in this exploratory setting. Future confirmatory studies are needed to further investigate the metabolite spectra of the identified *Faecalibacterium sp*. and *Oscillibacter sp*. 57_20.

Overall, the gut microbiome and metabolome exhibited a consistent pattern of fasting-induced alterations and nearly complete reversion to baseline at follow-up. Previous studies have consistently reported an enrichment of beneficial bacteria such as *F. prausnitzii* ^51-53^, as well as *Roseburia* and *Coprococcus* species, other known SCFA producers^54-56^. Our data only partly overlaps with these reports, as we observed no alterations to *F. prausnitzii*, alongside a decrease in *Roseburia sp*. and *Coprococcus sp*. However, we observed an increase in one *Butyricicoccus sp*., known butyrate producers. Our functional analysis revealed an increase in propionate production via kinase pathways, pointing towards an elevated SCFA production potential from coordinated activity across a broader microbial community rather than single species effects. The discrepancy with previous findings may be attributed to our study’s healthy cohort, in which baseline levels of *F. prausnitzii* are likely already well-balanced. This suggests that fasting might predominantly restore disturbed bacterial populations rather than altering already stable communities.^57^ Interestingly and consistent with previous studies, we did not observe an overall change in alpha diversity^5,55^. However, participants with lower baseline alpha diversity responded with a long-lasting increase to the fasting intervention. High microbial diversity stabilizes microbiota functions during perturbations^58^. In diseases usually associated with lower alpha diversity, such as obesity, other fasting interventions increase alpha diversity^59^. Together, these studies reinforce the hypothesis that fasting effects on alpha diversity are more pronounced in individuals with a reduced microbial diversity. However, low baseline alpha diversity may also arise from unaccounted external influences (e.g., lifestyle habits immediately preceding the measurement).

Despite these promising results, our study has several limitations. First, the lack of comprehensive documentation on dietary intake following the fasting intervention. Whether significant weight loss was solely due to fasting or also influenced by subsequent dietary modifications, a common limitation in caloric restriction studies cannot be determined. From a computational perspective, the relatively small sample size, particularly the initial test dataset after splitting, poses challenges for the random forest model and feature selection^60^. However, the testing of the classifier in three independent cohorts shows the robustness of our approach^61^.

In conclusion, our study provides compelling evidence that fasting induces substantial yet reversible changes in the gut microbiome and host metabolic profiles. Importantly, a subset of individuals experiences sustained weight loss. The integration of microbial and clinical data not only enhanced our predictive capabilities but also unveiled novel associations, such as the independent role of *Oscillibacter sp*. 57_20 and *Faecalibacterium sp*. incertae sedis in weight regulation. Despite limitations related to dietary documentation and sample size, these findings pave the way for future research aimed at refining personalized fasting interventions. Ultimately, our work underscores the promise of leveraging gut microbiome insights to tailor dietary strategies for improved metabolic health and weight management.

## Methods

### Study design

We performed a randomized, waitlist-controlled study (LEANER study, ClinicalTrials.gov; NCT04452916) carried out at the Experimental and Clinical Research Center of Charité – Universitätsmedizin in Berlin, from November 2020 to December 2022. The study protocol was approved by the institutional review board of Charité – Universitätsmedizin Berlin (EA1/033/20) and all participants provided written informed consent before entering the study. We collected and managed study data using REDCap electronic data capture tools hosted at Charité – Universitätsmedizin Berlin^62,63^. This study was exploratory, and we did not calculate a pre-determined sample size. Instead, the size of the recruited cohort was determined based on methodological and feasibility considerations. Flow diagram of recruitment and enrollment of patients according to Consort 2025 guidelines^64^ is shown in Figure S7. A general overview of the study is provided in Fig. 1a.

### Eligibility criteria

We included individuals aged 20-50 years who were assigned male or female at birth and identified as men and women, respectively; with a body mass index between 20-30 kg/m^2^. Exclusion criteria were heart, lung, liver, and kidney diseases requiring medical intervention, prescribed medication (except oral contraceptives), postoperative phases, current or chronic infection, antibiotic treatment or a fasting week within the last 6 months, habitual use of dietary supplements, food intolerances, > 2 kg body weight change within the last month, pregnancy, lactation, vegan diet, smoking, drug, or alcohol abuse.

### Intervention

Participants received oral and written instructions on the method of prolonged fasting. Expert guidance was provided by experienced nutritionists and certified fasting counselors in a group setting with 5-10 participants per group. After sufficient instruction, the fasting was performed individually at home, with the possibility of reaching out to the counselor at any time or to connect with other group members (if patients gave consent to share this information). Participants were encouraged but not required to take time off during this week. Fasting was preceded by one preparation day, on which participants consumed a low-calorie, fiber-rich diet, e.g. fruits, vegetables, potatoes, rice, and oats.

During the five fasting days, participants were allowed to consume 200-250 kcal/d through vegetable juices and vegetable broths. Herbal teas and water could be consumed ad libitum. At the morning of fasting day 6, we performed the study visit and patients were instructed for the refeeding phase. A detailed fasting plan including refeeding recommendation is provided as Supplementary Table S6.

### Randomization

We assigned participants randomly according to a locked list created by www.randomizer.at (two treatments: waiting, fasting; 1 factor: self-reported sex; permuted blocks; block sizes 4, 2, 6, 4). Due to the nature of the intervention and the wait-list design, blinding was not possible.

### Outcome measures

We established all outcome measures in advance and did not adapt them during or after the study. The primary outcome measure was resting energy expenditure (REE in kcal/d) after 5 days of fasting vs. baseline 1 assessed by indirect calorimetry. Secondary outcomes were body composition, cardiovascular parameters, various routine blood parameters, glucose variability, gut metagenome, fecal, and plasma metabolome. In addition, we assessed compliance and safety of fasting.

### Study center assessments

The waiting group underwent assessments four times, the fasting group three times. The fasting group had one baseline visit (baseline 1) and started the intervention within two to eleven days afterwards, the waiting group had two baseline visits with a 12-week waiting period in between (baseline 0 and baseline 1) and also started the intervention within a maximum of 11 days after baseline 1. These two baselines allowed detection of random time effects on the assessed outcome measures. For both groups, the next visit was on the morning of the sixth fasting day directly before breaking the fast (fasting). To assess medium-term effects of fasting, there was another visit after 12 weeks (follow-up). Except for the additional baseline 0 in the waiting group, all assessments were identical between the groups. All examinations described hereafter were performed at all study visits.

Assessments commenced in the morning following a 12-hour overnight fast. Participants were instructed to abstain from caffeine, alcohol, and vigorous exercise for 24 hours prior to the assessments.

Venous blood was drawn and sent to an accredited laboratory (Labor Berlin, Berlin, Germany) that measured parameters of glucose and lipid metabolism, blood count, electrolytes, liver enzymes, uric acid, CRP, and renal function according to international standards.

We measured waist and hip circumference to calculate waist-to-hip ratio and determined body composition (lean and fat mass) by air displacement plethysmography (Bod Pod, Life Measurements, Inc., Concord, CA).

Indirect calorimetry was conducted following a 30 min resting period while in a supine position within a thermoneutral environment for a duration of 30 min (Quark RMR, COSMED, Italy). REE and respiratory exchange ratio (RER) were calculated from oxygen consumption and carbon dioxide production as described elsewhere.^65^

Office blood pressure and heart rate were determined by five automated measurements within 10 min with an upper arm cuff (Mobil-o-Graph, I.E.M. GmbH, Stolberg, Germany) in the absence of study personnel.

### Fecal samples

At each study visit, participants received two stool collection kits and were asked to collect two samples from one stool at home as soon as possible following the study visit. One sample was used for gut metagenome sequencing (OMNIgene Gut, DNA Genotek, Ottawa, Canada) and the other for metabolome analysis (OMNImet Gut, DNA Genotek, Ottawa, Canada). These kits allow shipment without cooling to the study center. Participants sent the tubes to the study center at room temperature, where they were stored at - 80°C until further processing.

### Glucose sensors

We assessed glucose variability by continuous glucose monitoring over 14 days. For this, participants wore a glucose sensor that measured interstitial glucose concentrations in the subcutaneous tissue of the dorsal upper arm every 15 min (FreeStyle Libre Pro, Abbott, Chicago, IL, USA). In contrast to the use of these sensors in diabetic patients, participants in this study could not see their values during the measurement. Baseline assessment was done only in the waiting group directly following baseline visit 0. Additionally, fasting assessment was done in all participants during preparation, fasting, and refeeding.

### Ketone and glucose measurements

On the five fasting days, participants independently measured their blood concentration of beta-hydroxybutyrate (BHB) and glucose twice daily using a handheld glucometer (Freestyle Precision Neo, Abbott, Chicago, IL, USA), and recorded their values in a provided protocol. In addition, values were stored on the device and read out later in the study center, providing an objective measure of compliance.

### Fecal metagenomic sequencing

#### DNA isolation

DNA was isolated using ZymoBIOMICS DNA Miniprep Kit (ZymoReseach Europe GmbH, Freiburg, Germany). In short, an aliquot of 250 μL of stool sample from the OMNIgene Gut tube was added to a ZR BashingBead Lysis Tube. The following extraction was done according to the manufacturer’s instructions. DNA was eluted in 100 μL RNase/DNase-free water and collected in a 1.5-mL Eppendorf tube. Total DNA concentration was measured using NanoDrop ND-1000 (Peqlab Biotechnologie GmbH, Erlangen, Germany). Isolated DNA was stored at -80°C^66^.

#### Shotgun metagenomics

Isolated DNA was shipped on dry ice to Novogene (Cambridge, UK) for shotgun metagenomic DNA sequencing. In short, DNA was fragmented and subsequently underwent end repair and phosphorylation. After A-tailing, adapter ligation was performed. Then 150 bp paired-end sequencing was performed on the Illumina Novaseq 6000 platform with an aimed data collection of 6Gb raw data.

#### Microbiome data processing

Sequencing reads were mapped to the mOTUv2 (version 2.6) database, and read counts were subsequently rarefied using the RTK R package (version 0.2.6.1) to standardize sequencing depth across samples. Rarefaction involves randomly subsampling reads to match the sample with the lowest read depth, thereby mitigating biases introduced by variable sequencing depth and enabling reliable comparisons across cohorts. For the LEANER (healthy) cohort, a cutoff of 3,859 reads was applied, resulting in the exclusion of one sample due to insufficient sequencing depth. The same threshold was used for the Maifeld cohort^5^, where one sample was also excluded. In the Bahr dataset^22^, applying the same rarefaction criteria led to the exclusion of one sample. In the Ducarmon dataset^23^, applying the consistent rarefaction criteria resulted in the exclusion of 9 baseline samples (7 from GENESIS and 2 from OralFast trial). This consistent approach enabled reliable comparisons across cohorts and increased the robustness of downstream analyses.

### Plasma metabolomics

#### Collection of plasma

Venous blood was collected in Vacutainer® EDTA tubes (BD, Heidelberg, Germany) and immediately put on crushed ice for 30 min and then centrifuged (3,000 x g, 4°C, 10 min). Plasma was collected and stored at - 80°C.

#### Preparation of plasma for nuclear magnetic resonance spectroscopy (NMR)

Plasma samples were prepared with a robotic system (SamplePro Tube, Bruker BioSpin GmbH, Ettlingen, Germany) by mixing 287 μL plasma with 287 μL IVDr buffer (Bruker BioSpin GmbH, Ettlingen, Germany) and adding 10 μL formic acid (240 mM) as internal standard.

#### NMR measurement of plasma

All NMR experiments were performed on a 600 MHz Bruker Avance III spectrometer, using a triple resonance (^1^H, ^13^C, ^15^N, ^2^H lock) helium cooled cryoprobe with z-gradient. Samples were handled by an automatic Bruker SampleJet sample changer (Bruker Biospin GmbH, Ettlingen, Germany). Tuning and matching of the probe as well as locking and shimming of the sample were performed automatically. Following the Bruker IVDr protocol for plasma measurements for each sample, four different types of ^1^H NMR spectra were acquired at 310 K (1D NOESY, 2D JRES, 1D CPMG, and 1D spin echo diffusion spectrum).

#### Data analysis of plasma NMR spectra

From the collected spectra of each specimen, 41 smaller, non-lipoprotein metabolites were automatically identified and quantified using the Bruker IVDr Quantification in Plasma/Serum B.I.Quant-PSTM platform. Of note, only metabolites levels not bound to proteins were determined. Additionally, 112 lipoprotein parameters including various lipoprotein fractions, classes and subfractions were identified and quantified using the Bruker IVDr Lipoprotein Subclass Analysis B.I.LISATM platform.

### Stool metabolomics

#### Drying of stool samples

Fecal samples from OMNImet Gut tubes were processed for metabolomics. Fecal samples (1 mL) were dissociated two times after adding 2.5 mL 70% isopropanol using C tubes with the gentleMACS OctoDissociator (Miltenyi Biotec, Bergisch Gladbach, Germany). Aliquots of dissociated samples were covered with perforated Parafilm and dried overnight in a vacuum concentrator (Concentrator 5301, Eppendorf, Hamburg, Germany). Dried samples were stored at - 80°C.

#### Preparation of stool for NMR

Dried stool samples were resuspended in 600 μL (< 10 mg dry weight), 800 μL (10-40 mg dry weight) or 1,000 μL (> 40 mg dry weight) of double distilled water and 10 μL of an extraction standard (Nicotinic acid at 80 mmol/L) were added. Samples were centrifuged three times (12 000 x g, 4 °C, 10 min) and supernatants were transferred into a fresh vial after each centrifugation to remove particles. Of the final supernatant, 400 μL were mixed with 200 μL of 0.1 mol/L phosphate buffer (pH 7.4) and 50 μL of a 0.75% (w/v) solution of 3-trimethylsilyl-2,2,3,3-tetradeuteropropionate (TSP; Sigma-Aldrich, Taufkirchen, Germany), which served as an internal standard, in deuterium oxide.

#### NMR measurements of stool

NMR experiments were conducted on the same NMR spectrometer as described for the plasma analyses. Before measurement, each sample was allowed to equilibrate for 300 sec in the magnet, and the probe was automatically locked, tuned, matched, and shimmed. One -dimensional ^1^H NMR spectra were obtained at 298 K using a nuclear Overhauser enhancement spectroscopy pulse sequence with solvent signal suppression by pre-saturation during relaxation and mixing time. For each spectrum, 512 scans were collected into 65,536 data points over a 20-ppm (parts/million) spectral width using a relaxation delay of 4 sec, an acquisition time of 2.66 sec, and a mixing time of 0.01 sec. Spectra were automatically Fourier transformed and phase and baseline corrected.

#### Data analysis of stool NMR spectra

In each spectrum, a set of 41 metabolites were semi-automatically identified and relative to the TSP reference signal quantified using the CHENOMX 9.02 (Chenomx Inc, Edmonton, Canada) software suite. To account for potential losses during sample preparation, resulting data was adjusted to the extraction standard and normalized to dry weight.

### Predictive modelling of BMI response

To predict long-term BMI response to prolonged fasting, we used a random forest classifier implemented in the caret package (R 7.0.1). The outcome variable was responder status (responder vs. non-responder) defined by a BMI reduction exceeding the mean plus two standard deviations of the waiting list control group. Predictor variables included either (i) the full set of 87 microbiome and clinical features (Supplementary Data 6) or (ii) a predefined sparse model with four predictors: *Oscillibacter sp*. 57_20, *Faecalibacterium sp*. incertae sedis, systolic blood pressure, and LDL cholesterol. Given the limited sample size (n = 34), we used 100 stratified 80/20 Monte-Carlo resamples with inner 5-fold cross-validation for hyperparameter tuning, using the area under the ROC curve (AUC) as the optimization metric. The best-performing model from each resample was evaluated on its holdout 20% test partition, providing a distribution of performance estimates rather than a single value. Model performance was assessed by AUC, accuracy, sensitivity, specificity, balanced accuracy, F1 score, Matthews correlation coefficient (MCC), and average precision (area under the precision–recall curve). We report both mean ± SD and median [IQR] across resamples to capture variability. Feature importance was obtained via permutation importance, and stability summarized as mean importance ± SD and rank distribution. Feature-stability analysis showed *Oscillibacter sp*. 57_20 and *Faecalibacterium sp*. incertae sedis consistently ranked highest, systolic BP contributed modestly, and LDL showed low and variable importance. LDL was retained in the predefined four-feature model to avoid post-hoc selection, while a three-feature variant performed comparably (Supplementary Table S3).

To assess potential confounding and the added predictive value of microbiome data, we trained two additional models using the same resampling procedure: (i) a clinical-only model (age, sex, baseline BMI, blood pressure, lipid parameters), and (ii) a combined model incorporating microbiome predictors.

### Calibration

To evaluate the reliability of predicted probabilities, we applied post-hoc calibration using Platt scaling (logistic correction of the logit scores) and isotonic regression (monotone, non-parametric mapping) in the Maifeld and Ducarmon cohort. The original model was not retrained. Instead, we performed 100 stratified 80/20 Monte-Carlo resamples, ensuring that both outcome classes were present in each training and test set. For every resample, the calibration models were fit on the training fold only and then applied to the hold-out test fold. We compared three variants of prediction: the raw model (no calibration), Platt-scaled, and isotonic-scaled probabilities.

On each test fold, we calculated AUC, accuracy, sensitivity, specificity, balanced accuracy, F1 score, Matthews correlation coefficient (MCC), and the Brier score (mean squared error of predicted probabilities). Predictions were dichotomized at a 0.5 threshold to form confusion matrices, summarized as mean ± SD of the test proportion. Calibration was further assessed using Calibration-in-the-Large (CITL) and Calibration Slope in the test fold. All performance measures were summarized as mean ± SD across the 100 repeats.

### Statistical analysis

Descriptive statistics for interval variables are presented as mean ± standard deviation or median with interquartile range (IQR). For comparisons of delta BMI between responders and non-responders, normality of variable distributions was assessed by visual check of histograms and qq-plots. Mann-Whitney U-test or t-test was used as appropriate. For all other data, comparisons between independent groups were performed using the Mann-Whitney U test, and paired samples were compared using the Wilcoxon signed-rank test. Where applicable, p-values were adjusted for multiple comparisons using either the Holm method (for family-wise error control) or the Benjamini-Hochberg (BH) procedure (for false discovery rate control), depending on the analytical objective.

Permutational Multivariate Analysis of Variance (PERMANOVA) was used to test for multivariate compositional differences between groups and visits.

To evaluate the relationship between bacterial abundance and metabolite concentrations while accounting for visit timepoint, we performed permutation-based conditional independence tests. Specifically, for each metabolite, its association with *Oscillibacter* sp. 57_20 and *Faecalibacterium* sp. incertae sedis was tested while conditioning on the visit variable, using the R package coin (version 1.4.3) with 10,000 permutations. Resulting p-values were corrected for multiple testing using the Benjamini-Hochberg false discovery rate (FDR) method.

Statistical significance was defined as p < 0.05 or q < 0.1, where applicable.

## Data Availability

All code, data, and scripts used in this study have been made publicly available. The code, scripts and processed raw data can be accessed on Zenodo (doi: https://doi.org/10.5281/zenodo.15696891). Raw sequencing data will be deposited in the NCBI database upon full publication.

## Data and code availability

All code, data, and scripts used in this study have been made publicly available. The code, scripts, and processed raw data can be accessed on Zenodo (doi: https://doi.org/10.5281/zenodo.17493808). Raw sequencing data will be made available under the BioProject ID PRJNA1293883 from the NIH Sequence Read Archive.

## Acknowledgement

We would like to thank Anja Mähler for designing and setting up the LEANER study, for recruiting the study cohort, the entire study organization, and establishing the network around this fasting study. We thank Alexandra Prüß who assisted as a fasting guide in carrying out study protocols. We thank Jana Czychi for her excellent technical assistance. This work was funded by intramural funds of the Experimental and Clinical Research Center, a cooperation between the Max Delbrück Center for Molecular Medicine in the Helmholtz Association and Charité – Universitätsmedizin Berlin; and external funds from the Karl and Veronica Carstens foundation (KVC 0/120/2021).

FP, NW, HB, DNM, WG and PJO were supported by the BMFTR (Federal Ministry of Research, Technology and Space), project IDs 01EJ2202A, 01EJ2202D and 01EJ2202B (TAhRget consortium). FP, NW, and SKF were supported by the European Union (HORIZON-HLTH-2022-STAYHLTH-02, IMMEDIATE consortium). TUPB was supported by the Berlin Center for Translational Vascular Biomedicine, jointly founded by BIH, Charité - Universitätsmedizin, and the Max Delbrück Center. DNM and NW were supported by the DZGIF (DZG Innovation Fund), Topic “Microbiome”. SKF, DNM and NW were supported by the German Research Foundation (DFG), project ID 437531118 (CRC1470, projects A05, A06 and A10). SKF was supported by the DFG project TRR412 and DZHK. AZ was supported by the DFG, project ID 324392634 (TR221).

## Author Contributions

CRediT: Conceptualization: TUPB, SB, HB, NS; Data curation: GNK, TUPB, LSB, FG, QRD, GZ, RM, SB, HB; Formal Analysis: GNK, TUPB, UL, MM, AK, HB; Funding acquisition: TUPB, NW, PJO, WG, DNM, SKF, SB, HB, NS; Investigation: KK, TS, SK, GR, VM, LB, LM, LSB, FG, FP, QRD, GZ, RM, SB, NS; Methodology: GNK, TUPB, UL, WG, PJO, SKF, HB; Project administration: SB, HB, NS; Resources: NW, AZ, PJO, WG, DNM, SKF, SB, HB, NS; Software: GNK, TUPB, UL, AK, SKF; Supervision: TUPB, UL, AZ, PJO, WG, DNM, SKF, SB, HB, NS; Validation: GNK, TUPB, LSB, FG, FP, QRD, GZ, RM, HB; Visualization: GNK, TUPB, HB; Writing – original draft: GNK, TUPB, SB, HB, NS; Writing – review & editing: all authors.

## Conflict of interest

NS has received speaker honoraria from Teva GmbH, Alexion Pharma Germany GmbH and Roche Pharma AG. RM is employed by the Buchinger Wilhelmi Development & Holding GmbH. The other authors declare no competing interests.

## Notes

### Competing Interest Statement

NS has received speaker honoraria from Teva GmbH, Alexion Pharma Germany GmbH and Roche Pharma AG. The other authors declare no competing interests.

### Clinical Trial

NCT04452916

### Funding Statement

This study was funded by intramural funds of the Experimental and Clinical Research Center, a cooperation between the Max Delbruck Center for Molecular Medicine in the Helmholtz Association and Charite Universitatsmedizin Berlin; and external funds from the Karl and Veronica Carstens foundation (KVC 0/120/2021).

### Author Declarations

The study carried out at the Experimental and Clinical Research Center of Charite Universitatsmedizin Berlin, Berlin, Germany, from November 2020 to December 2022. The study protocol was approved by the institutional review board of Charite Universitatsmedizin Berlin (EA1/033/20) and all participants provided written informed consent before entering the study.

### Summary of Updates

Inclusion of a third cohort for application of the machine learning model. Improved characterisation of the predictive model. Correction of minor mistakes (e.g., spelling).

