## Supplementary Methods, Figures, and Tables for "Machine Learning Identifies Microbiome and Clinical Predictors of Sustained Weight Loss Following Prolonged Fasting"

Kaufhold*, Bartolomaeus*, et al.

**Supplementary Tables**

**Table S1.** Fasting-related adverse events.

| **Adverse event** | **n (%)** |
| --- | --- |
| Circulatory dysregulation | 10 (26.3%) |
| Weakness, fatigue | 5 (13.2%) |
| Freezing | 3 (7.9%) |
| Nausea, vomiting | 2 (5.3%) |
| Sleep disturbance | 2 (5.3%) |
| Back pain | 2 (5.3%) |
| Leg muscle pain | 2 (5.3%) |

**Table S2:** Feature contributions and model performance of the short 4-feature model.

**A)** Mean feature importance and rank stability across 100 repeated stratified 80/20 resamples, indicating the relative contribution of each predictor to model performance. ldl, low-density lipoprotein: SD, standard deviation; sys_mean, systolic blood pressure.

| **Feature** | **Importance (mean ± SD)** | **Rank (median [IQR])** |
| --- | --- | --- |
| Oscillibacter.sp..57_20. | 13.09 ± 5.93 | 1 [1–2] |
| NA.Faecalibacterium.species.incertae.sedis | 11.01 ± 3.98 | 2 [1–2] |
| sys_mean | 5.27 ± 2.72 | 3 [3–3] |
| ldl | -0.94 ± 3.16 | 4 [4–4] |

**Table S3.** Comparison of predictive performance of the short model with and without LDL: Predictive performance of the short model including all four features compared with a three-feature variant excluding LDL. Values represent mean ± SD across 100 repeated stratified 80/20 resamples.AUC, area under the receiver operating characteristic curve; SD, standard deviation; F1, F1-score; MCC, Matthews correlation coefficient; LDL, low-density lipoprotein.

| **Metric** | **Short (4 features with LDL)** | **Short (3 feats, w/o LDL)** |
| --- | --- | --- |
| AUC | 0.86 ± 0.16 | 0.91 ± 0.13 |
| Accuracy | 0.79 ± 0.15 | 0.82 ± 0.14 |
| Sensitivity | 0.65 ± 0.34 | 0.74 ± 0.29 |
| Specificity | 0.86 ± 0.17 | 0.86 ± 0.17 |
| Balanced Accuracy | 0.75 ± 0.18 | 0.80 ± 0.16 |
| F1 | 0.73 ± 0.19 | 0.75 ± 0.18 |
| MCC | 0.57 ± 0.35 | 0.64 ± 0.30 |

**Table S4. Calibration for the Maifeld cohort.** To panel: Mean ± SD predictive performance metrics (AUC, Accuracy, Sensitivity, Specificity, Balanced Accuracy, F1, and MCC) across 100 stratified 80/20 train–test resamples. Recalibration was performed using Platt scaling and isotonic regression on the training folds only and evaluated on the held-out test folds. The raw model (no recalibration) is shown for comparison under the same split strategy. Middle panel: Calibration-in-the-large (CITL) and calibration slope (Slope) values (Mean ± SD). Lower panel: Confusion matrix metrics (% of total test data) illustrating Mean ± SD true/false positives and negatives and resulting accuracy across repetitions.

| **Prediction of responder status** | | | | | | | | | | | |
| --- | --- | --- | --- | --- | --- | --- | --- | --- | --- | --- | --- |
| **Method** | **AUC**  **mean ± SD** | **Accuracy**  **mean ± SD** | **Sensitivity**  **mean ± SD** | **Specificity**  **mean ± SD** | **Balanced Accuracy**  **mean ± SD** | | | | **F1**  **mean ± SD** | **MCC**  **mean ± SD** | **Brier**  **mean ± SD** |
| **isotonic** | 0.793 ± 0.139 | 0.740 ± 0.133 | 0.790 ± 0.180 | 0.640 ± 0.318 | 0.715 ± 0.156 | | | | 0.794 ± 0.120 | 0.451 ± 0.330 | 0.201 ± 0.097 |
| **platt** | 0.843 ± 0.136 | 0.667 ± 0.109 | 0.853 ± 0.155 | 0.295 ± 0.319 | 0.574 ± 0.141 | | | | 0.769 ± 0.085 | 0.156 ± 0.328 | 0.205 ± 0.066 |
| **raw** | 0.843 ± 0.136 | 0.818 ± 0.119 | 0.785 ± 0.159 | 0.885 ± 0.211 | 0.835 ± 0.123 | | | | 0.845 ± 0.110 | 0.661 ± 0.235 | 0.203 ± 0.039 |
| **Calibration-in-the-large and calibration slope** | | | | | | | | | | |  |
|  | **CITL**  **mean ± SD** | **p value (CITL)** | **Slope**  **mean ± SD** | **p value (Slope)** | | | **Note** | | | |  |
| **Isotonic** | NA | NA | NA | NA | | | Not possible for isotonic regression. | | | |  |
| **platt** | -0.417 ± 0.953 | 0.661 | 0.297 ±  0.948 | 0.458 | | | Ok | | | |  |
| **raw** | 0.497 ± 0.883 | 0.573 | 0.657±  2.100 | 0.870 | | | Ok | | | |  |
| **Confusion matrix** | | | | | | | | | | |  |
|  | **TP % test data** | **FP % test data** | **TN % test data** | **FN % test data** | | **Accuracy % test data** | |  | | | |
| **isotonic** | 52.667 ± 12.014 | 12.000 ± 10.615 | 21.333 ± 10.615 | 14.000 ± 12.014 | | 74.000 ± 13.257 | |  | | | |
| **platt** | 56.833 ± 10.352 | 23.500 ± 10.619 | 9.833 ± 10.619 | 9.833 ± 10.352 | | 66.667 ± 10.856 | |  | | | |
| **raw** | 52.333 ± 10.599 | 3.833 ± 7.049 | 29.500 ± 7.049 | 14.333 ± 10.599 | | 81.833 ± 11.867 | |  | | | |

**Table S5. Calibration for the Ducarmon cohort.** To panel: Mean ± SD predictive performance metrics (AUC, Accuracy, Sensitivity, Specificity, Balanced Accuracy, F1, and MCC) across 100 stratified 80/20 train–test resamples. Recalibration was performed using Platt scaling and isotonic regression on the training folds only and evaluated on the held-out test folds. The raw model (no recalibration) is shown for comparison under the same split strategy. Middle panel: Calibration-in-the-large (CITL) and calibration slope (Slope) values (Mean ± SD). Lower panel: Confusion matrix metrics (% of total test data) illustrating Mean ± SD true/false positives and negatives and resulting accuracy across repetitions.

| **Prediction of responder status** | | | | | | | | | | |
| --- | --- | --- | --- | --- | --- | --- | --- | --- | --- | --- |
| **Method** | **AUC mean ± SD** | **Accuracy**  **mean ± SD** | **Sensitivity**  **mean ± SD** | **Specificity**  **mean ± SD** | | **Balanced Accuracy**  **mean ± SD** | | **F1**  **mean ± SD** | **MCC**  **mean ± SD** | **Brier**  **mean ± SD** |
| **isotonic** | 0.625 ± 0.117 | 0.529 ± 0.104 | 0.645 ± 0.249 | 0.390 ± 0.276 | | 0.517 ± 0.104 | | 0.578 ± 0.149 | 0.030 ± 0.233 | 0.269 ± 0.060 |
| **platt** | 0.679 ± 0.146 | 0.573 ± 0.104 | 0.797 ± 0.196 | 0.304 ± 0.179 | | 0.550 ± 0.101 | | 0.661 ± 0.114 | 0.130 ± 0.247 | 0.243 ± 0.037 |
| **raw** | 0.679 ± 0.146 | 0.605 ± 0.125 | 0.658 ± 0.168 | 0.542 ± 0.185 | | 0.600 ± 0.125 | | 0.640 ± 0.129 | 0.209 ± 0.258 | 0.242 ± 0.056 |
| **Calibration-in-the-large and calibration slope** | | | | | | | | | |  |
|  | **CITL**  **mean ± SD** | **p value (CITL)** | **Slope**  **mean ± SD** | **p value (Slope)** | **Note** | | | | |  |
| **Isotonic** | NA | NA | NA | NA | Not possible for isotonic regression. | | | | |  |
| **platt** | 0.163 ± 0.753 | 0.784 | 2.290 ±  1.970 | 0.512 | Ok | | | | |  |
| **raw** | 0.843 ± 0.263 | 0.263 | 0.493 ±  0.424 | 0.232 | Ok | | | | |  |
| **Confusion matrix** | | | | | | | | | |  |
|  | **TP % test data** | **FP % test data** | **TN % test data** | **FN % test data** | **Accuracy % test data** | |  | | | |
| **isotonic** | 35.182 ± 13.592 | 27.727 ± 12.553 | 17.727 ± 12.553 | 19.364 ± 13.592 | 52.909 ± 10.368 | |  | | | |
| **platt** | 43.455 ± 10.700 | 31.636 ± 8.119 | 3.818 ± 8.119 | 11.091 ± 10.700 | 57.273 ± 10.377 | |  | | | |
| **raw** | 35.909 ± 9.171 | 20.818 ± 8.403 | 24.636 ± 8.403 | 18.636 ± 9.171 | 60.545 ± 12.476 | |  | | | |

**Table S6.** Patient information for the fasting Intervention and recommendations.

| **Day** | **Notes** | **Food Plan** | **Optional and additional recommendations** | **Exercise & Relaxation** |
| --- | --- | --- | --- | --- |
| **Day 0: Preparation** |  | Preparation of fasting foods, beverages, and a mental approach to fasting. | Fibre-rich foods. | Outdoor movement, relaxation, and detachment from daily routine. |
| **Day 1: Fasting** | Morning: Laxative measures (Glauber's salt) | Morning: 150 ml vegetable juice  Afternoon: 150 ml vegetable juice  Evening: Vegetable broth | Liver compresses.  Enema. | Light outdoor movement, supportive exercises, rest periods. |
| **Day 2-5: Continued Fasting** |  | Morning: 150 ml vegetable juice  Afternoon: 150 ml vegetable juice  Evening: Vegetable broth | Liver compresses.  Enema. | Outdoor activity, balanced with rest phases. Avoid strenuous effort. |
| **Day 6: Refeeding Day** | Fasting examination (Fasting visit) in the morning and afterward re-introduction of food | Morning: Oatmeal with apple or berries and cinnamon  Afternoon: Potato-vegetable soup or vegetable dish  Evening: Fresh vegetable plate or grain-based dish | Liver compresses. | Engage in mild physical activity, continue relaxation practices. |
| **Day 7: Refeeding Days** |  | Continue reintroducing vegetables, grains, and proteins gradually. | Slow increase in grain intake, add fats on Day 7, proteins on Day 8. Maintain low salt. | Regular outdoor activity, maintain rest periods. |

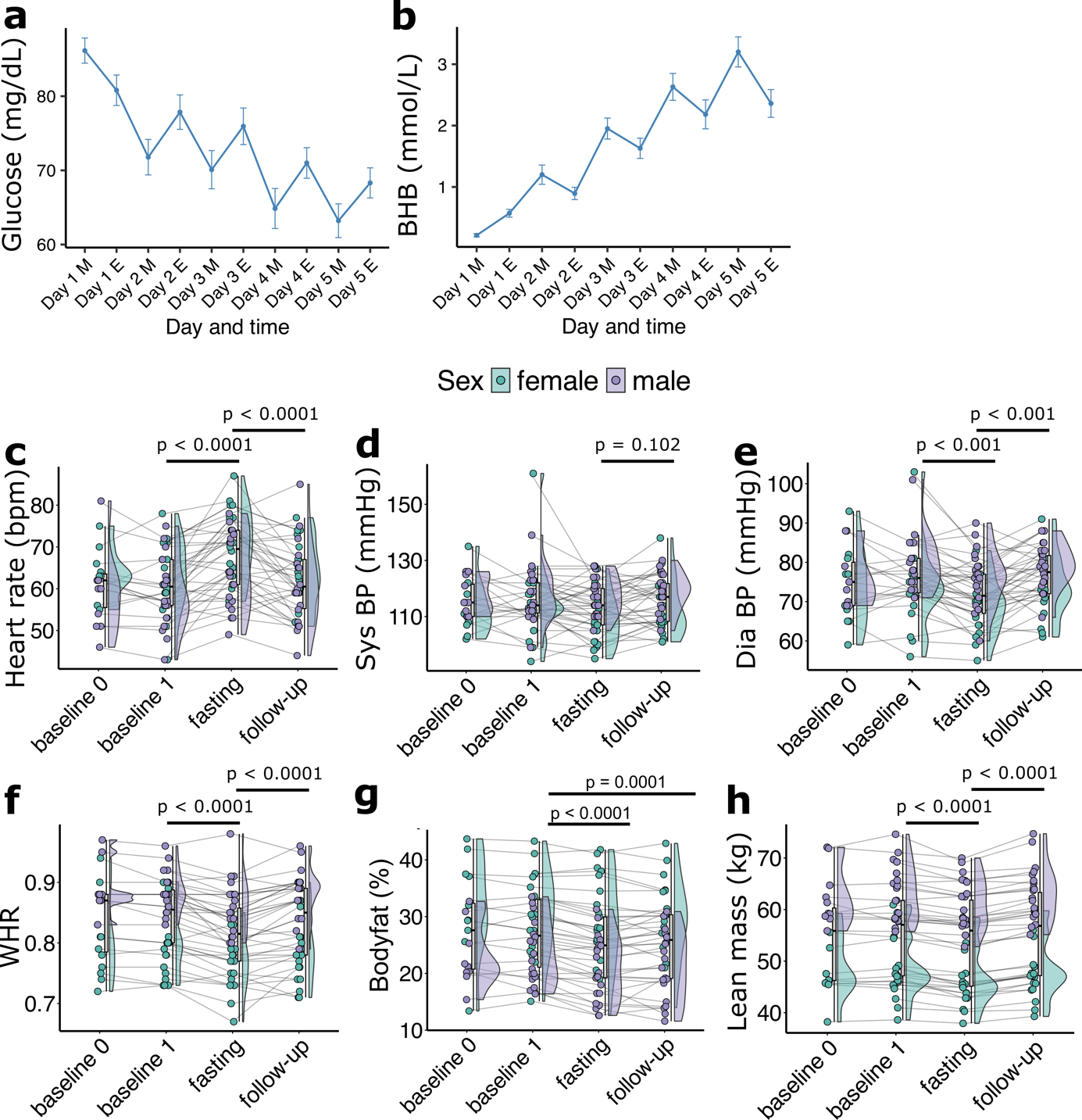

**Figure S1.** **A)** Glucose levels and **B)** BHB (beta-hydroxybutyrate) levels measured during days 1 to 5 of fasting, shown as a time series for both morning and evening measurements. **C)** Heart rate (bpm), **D)** Systolic blood pressure (Sys BP, mmHg), **E)** Diastolic blood pressure (Dia BP, mmHg), **F)** Waist-to-hip ratio (WHR), **G)** relative body fat, and **H)** Lean mass across the study visits (baseline, fasting, and follow-up visits). For panels **D-H)**, each point represents one participant, with lines connecting individual measurements across timepoints. The middle boxplots represent all participants, while violins on the right show the distribution of values. Data points and violines are colored by sex. Q-values for significance were determined using Wilcoxon signed-rank tests with Benjamini-Hochberg false discovery rate (BH-FDR) correction, with significance indicated as q < 0.1, q < 0.01, and q < 0.001.

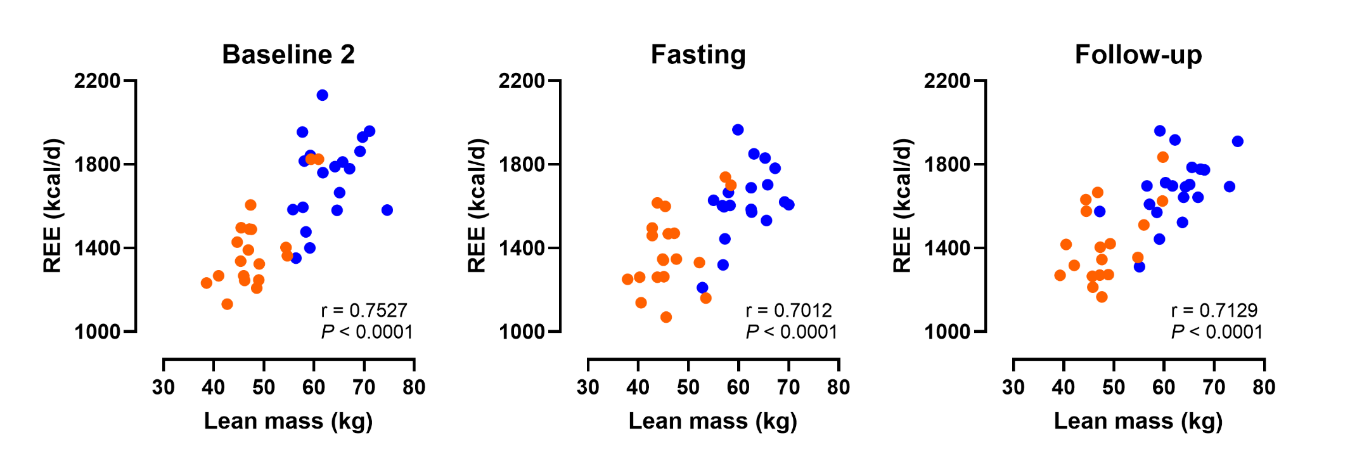

**Figure S2.** Pearson's correlation between Lean mass and resting energy expenditure across the study visits (baseline, fasting, and follow-up visits) stratified by sex (female = orange, male = blue).

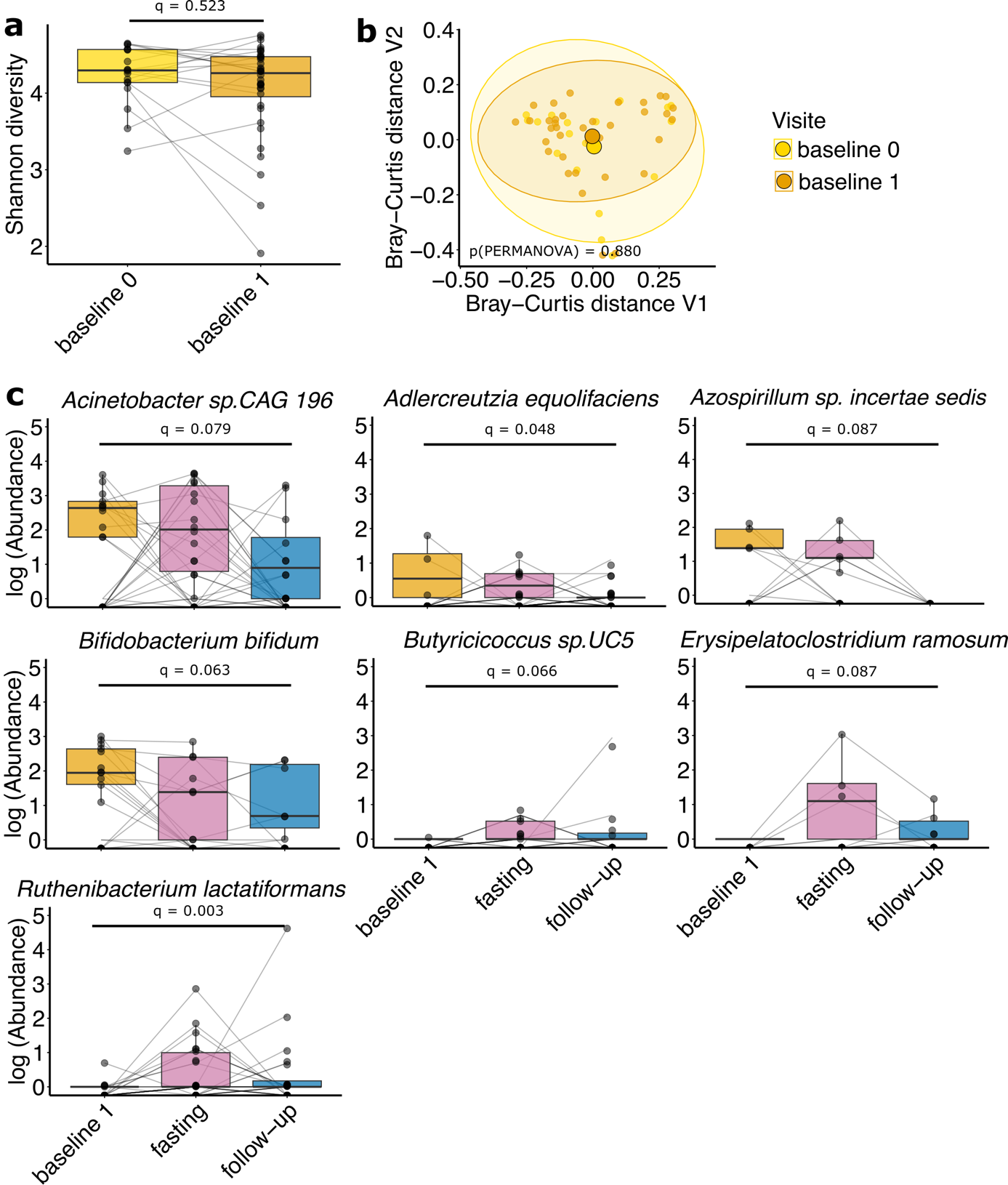

**Figure S3. A)** Alpha diversity (Shannon index) for both baseline visits (baseline 0 and 1). **B)** Principal coordinate analysis (PCoA) based on Bray–Curtis dissimilarity for both baseline visits. Ellipses represent 95% confidence intervals p-values from PERMANOVA. **C)** Microbial species (taxonomic level) significant different between baseline1 and follow-up. Boxplots coloured by visit. Significance by Wilcoxon signed-rank test with Benjamini–Hochberg false discovery rate correction.

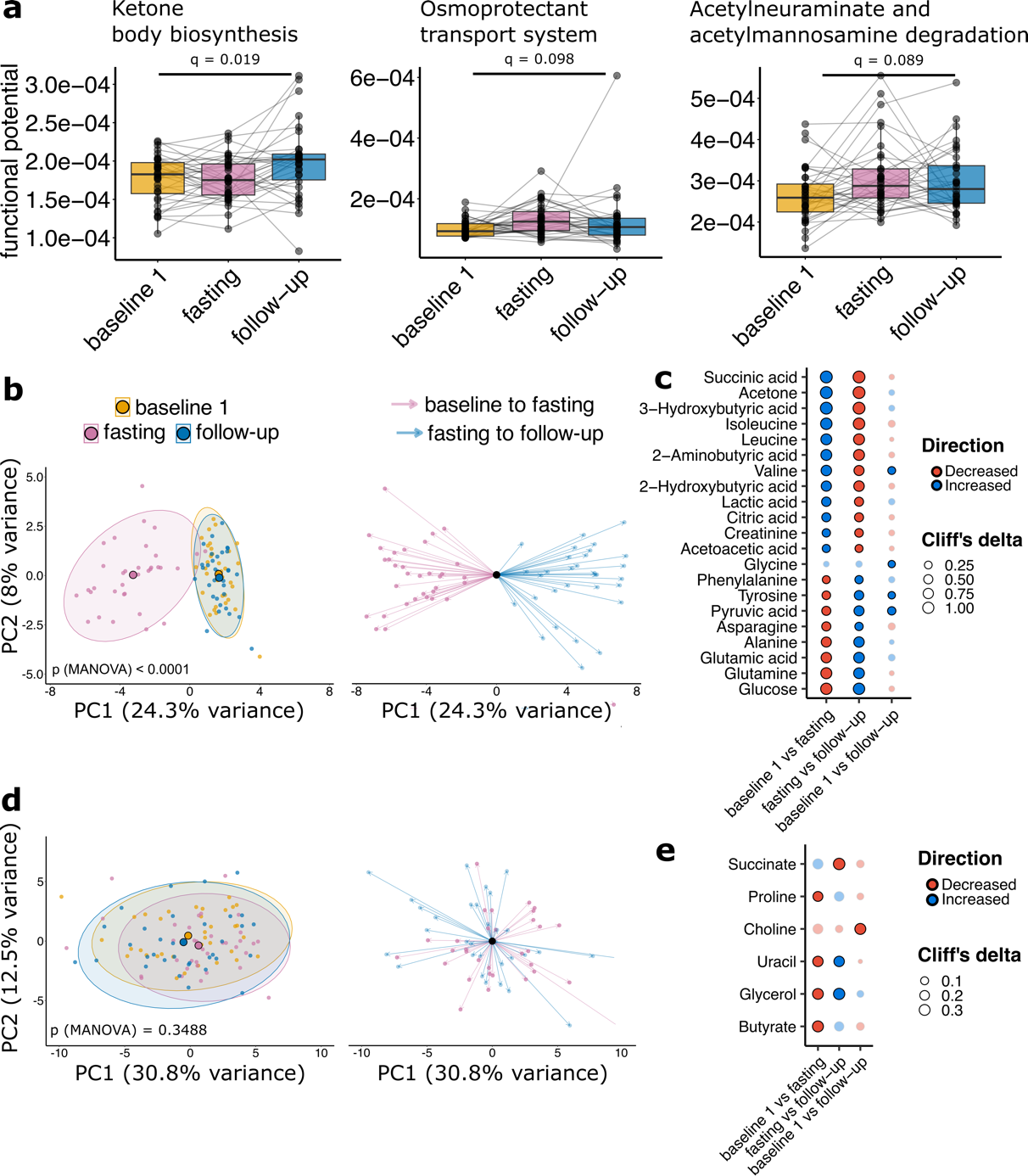

**Figure S4. A)** Gut metabolic modules significant different between baseline1 and follow-up. Boxplots coloured by visit. Significance by Wilcoxon signed-rank test with Benjamini–Hochberg false discovery rate correction. **B)** Plasma metabolome and **D)** stool metabolome principal coordinate analysis (PCA) (naïve, left) and centered on baseline visit for fasting effect and on fasting visit for post-fasting effect (right). Ellipses represent 95% confidence intervals p-values from PERMANOVA. Dot plots showing differential abundant **C)** plasma metabolites and **E)** stool metabolites. Dots show fasting effects, follow-up effects and study effects, transparency indicates non-significant findings (q > 0.1), dot size shows absolute Cliff’s delta, color shows directionality. Significance by Wilcoxon signed-rank test with Benjamini–Hochberg false discovery rate correction.

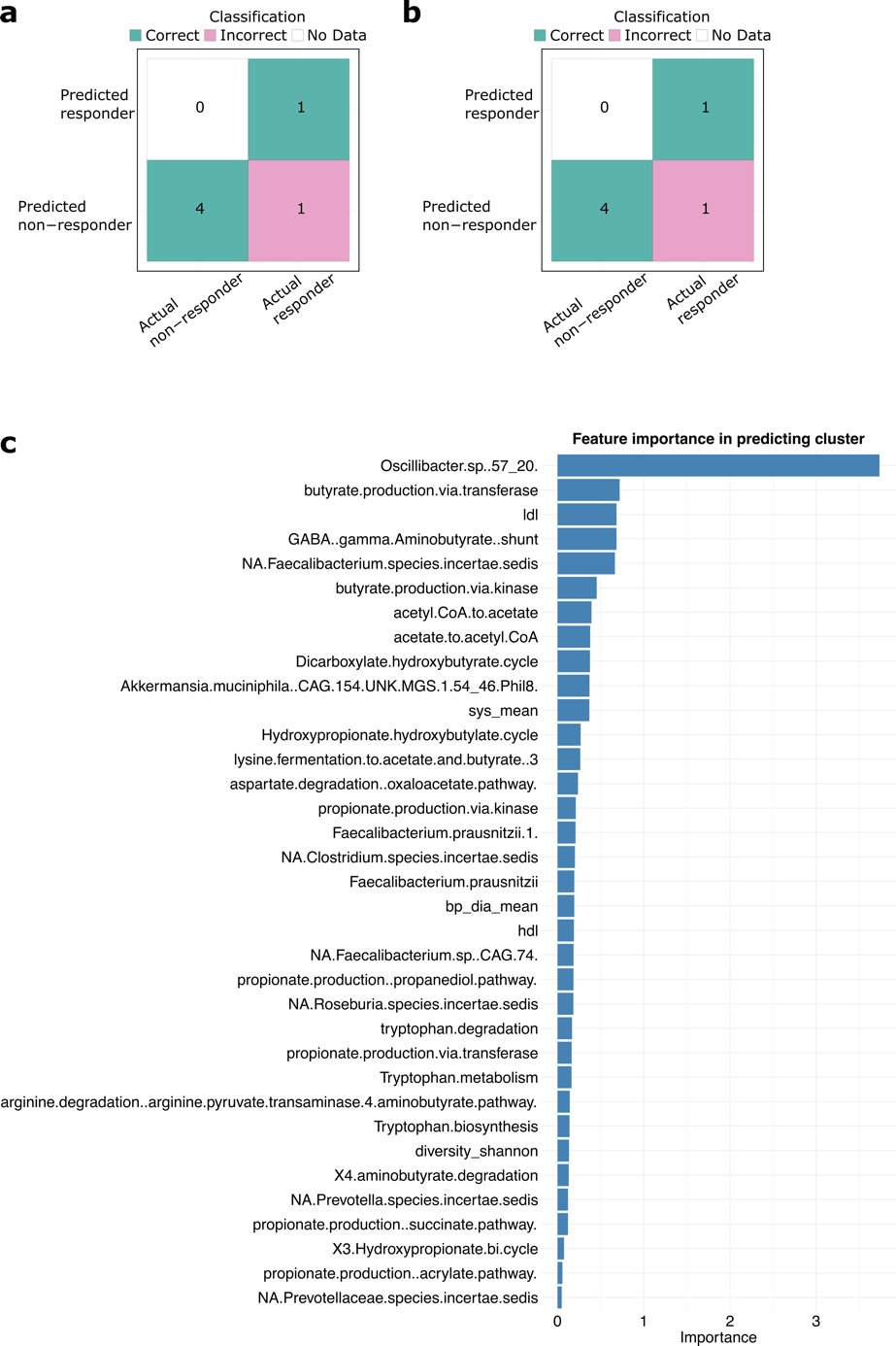

**Figure S5.** Confusion matrix of the model tested in 20% test data **A)** without and **B)** with BMI. **C)** Models feature importance, showing the relative contributions of each feature in predicting the outcome.

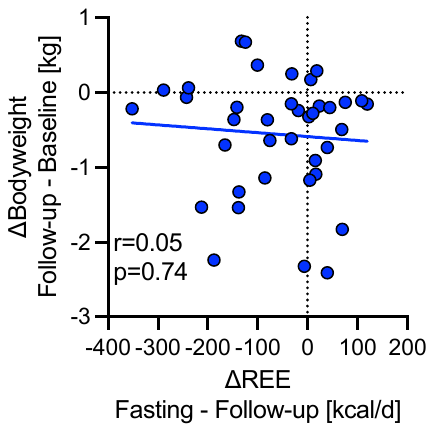

**Figure S6.** The correlation between delta body weight (Follow-up – Baseline) and delta REE (Fasting – Follow-up) is shown.

Discontinued intervention (n = 0)

Lost to follow-up for primary outcome (no response at V4) (n = 1)

Excluded (n = 7)

Not meeting inclusion criteria (n = 7)

Declined to participate (n = 0)

Other reasons (n = 0)

Analysis

Analysed for primary outcome (n = 19)

Excluded from analysis (n = 0)

Discontinued intervention (n = 0)

Lost to follow-up for primary outcome (n = 0)

Randomised (n = 55)

Allocation

Follow-Up

Allocated to Fasting (n = 29)

Received allocated intervention (n = 19)

Did not receive allocated intervention (n=10) ((SARS-CoV2 infection (n=1), (pregnancy (n=1), antibiotic treatment (n=1), declined to participate (n=7))

Allocated to Waiting (n = 26)

Received allocated intervention (n = 20)

Did not receive allocated intervention (n=6) ((Corona infection (n=1), rehabilitation after surgery (n=1), antibiotic treatment (n=1), declined to participate (n=1), no response (n=2))

Enrolment

Assessed for eligibility (n = 62 )

Analysed for primary outcome (n = 19)

Excluded from analysis (n = 0)

**Figure S7.** CONSORT 2025 Flow Diagram. Flow diagram of the progress through the phases of a randomised trial of two groups (that is, enrolment, intervention allocation, follow-up, and data analysis).
